# An Evolutionary Perspective on the Genetics of Anorexia Nervosa

**DOI:** 10.1101/2023.08.09.23293879

**Authors:** Édith Breton, Tobias Kaufmann

## Abstract

Anorexia nervosa (AN) typically emerges around adolescence and predominantly affects females. Recent progress has been made in identifying biological correlates of AN, but more research is needed to pinpoint the specific mechanisms that lead to its development and maintenance. There is a known phenotypic link between AN, growth and sexual maturation, yet the genetic overlap between these phenotypes remains enigmatic. One may hypothesize that shared factors between AN, energy metabolism and reproductive functions may have been under recent evolutionary selection. Here, we characterize the genetic overlap between AN, BMI and age at menarche, and aimed to reveal recent evolutionary factors that may help explain the origin of AN. We obtained publicly available GWAS summary statistics of AN, BMI and age at menarche and studied the polygenic overlap between them. Next, we used Neandertal Selective Sweep scores to explore recent evolutionary selection. We found 22 loci overlapping between AN and BMI, and 9 loci between AN and age at menarche, with 7 of these not previously associated with AN. We found that loci associated with AN may have been under particular evolutionary dynamic. Chronobiology appeared relevant to the studied genetic overlaps and prone to recent evolutionary selection, offering a promising avenue for future research. Taken together, our findings contribute to the understanding of the genetic underpinning of AN. Ultimately, better knowledge of the biological origins of AN may help to target specific biological processes and facilitate early intervention in individuals who are most at risk.

## Introduction

Anorexia Nervosa (AN) is a serious psychiatric disorder with long-lasting consequences for physical health, psychological well-being and quality of life (1–3). Symptoms of AN typically include severe restriction of energy intake, intense fear of gaining weight, and disturbed body image (2). Notably, research on AN and other eating disorders (ED) has lagged significantly behind that of other major psychiatric disorders (2,4), particularly with regard to the biological mechanisms involved in their development. Yet, AN has one of the highest mortality and morbidity rates of all psychiatric disorders – with a standardized mortality ratio of 5.9 (5).

Restrictive EDs affect approximately one male for every 10 females, and commonly emerge around puberty, with a median age at onset of 12 years (6). Major neurodevelopmental changes occur during this period, potentially increasing the vulnerability to develop a mental disorder (7). Growth may also play a role, since children with both lower and higher body mass index (BMI) before adolescence may be at increased risk of EDs (8–11).

Earlier age at onset and time spent with an ED are predictors of poorer outcomes, supporting the need to intervene in the early stages of the illness (2,3,12,13). Evidence from longitudinal studies suggests that ∼30% of individuals with AN will recover in the first 10 years after diagnosis, and that an additional ∼30% will recover after 20 years (1,14,15), suggesting fair chances for recovery. Yet, a third of individual with AN will still be ill after 20 years, and a large proportion will live with psychiatric symptoms for a significant part of their lives, even after remission from their ED (14,16).

Understanding the genetic underpinnings of AN could provide an opportunity to identify biological mechanisms at play – which could eventually become targets for improving interventions. Yet, studying the genetic origins of AN is challenging, in part due to its early age at onset and sexes imbalance, but perhaps also because people with AN often do not seek help for their condition (4,15), adding to the difficulty of obtaining sufficient sample sizes for genome-wide analyses (GWAS). A recent AN GWAS identified eight risk loci and displayed significant pleiotropy with several psychiatric and metabolic traits (17). Further, they showed that AN is genetically correlated with several psychiatric and metabolic traits. The study also provided evidence that low weight may not only be a consequence of AN-related cognitions, but that some metabolic traits predisposing to low weight may facilitate the development and maintenance of AN (17,18).

An aspect that is often overlooked in the study of AN is evolution. The heritability estimate of 48-74% (19) may suggest that some AN-associated genes could have been evolutionary advantageous, for example in terms of nutrition and reproduction (20–22). For example, one may hypothesize that at some point in recent human evolution, it may have been beneficial to be able to stay physically active in order to travel and forage continuously in situations where food was scarce, and some genes associated with AN may have conferred some metabolic advantage in this regard (20–22).

An evolutionary perspective on the genetics of AN may also provide a better understanding of the link between AN and puberty. Since periods of growth such as childhood and adolescence are energy demanding, one may hypothesize that individuals who matured more quickly may have become less demanding more quickly in terms of resource use, benefiting their group. Because AN is often associated with amenorrhea, some authors have proposed that AN might have been a way to supress reproductive capacity when environmental conditions were not favourable, in order to increase reproductive success by saving energy for more optimal conditions (20–22). This would be consistent with results from studies assessing the link between AN and sexual maturation, which indicate possible delay in individuals with AN (23,24), although others have found evidence that may suggest otherwise (25). It is also possible that genes impacting puberty in either direction may have been beneficial in terms of survival and reproduction depending on environmental scenarios (26,27), yet those genes might also modulate the risk of AN in today’s living condition.

One way to study such evolutionary adaptations is to compare the genome of modern humans (Homo sapiens) to an extinct, close relative – *Homo neanderthalensis* – which might reveal *sapiens*-specific features that may have provided our specie with an evolutionary advantage (28). Similar approaches have recently implied the role of recent evolution in the genetics of schizophrenia (29).

The overarching aims of the present study were to (1) deepen knowledge of the genetic architecture of AN through its associations with BMI and age at menarche, and to identify novel loci shared between those traits, and (2) to gain a better understanding of the evolutionary origins of AN in general, and the identified loci in particular.

## Methods

### Samples

The present study uses publicly available summary statistics from GWAS of AN (17), BMI (30), and age at menarche (31) (see supplements for details). All GWAS included individuals predominantly of European ancestry. Data collection for each GWAS was performed with participants’ written informed consent and with approval by the respective local Institutional Review Boards, as specified in the respective studies (17,30,31).

### Statistical Analyses

#### Genetic correlation

We calculated genetic correlations between AN and BMI, as well as between AN and age at menarche, using LD-score regression (v1.0.1), available at https://github.com/bulik/ldsc. These genetic correlations have been reported in previous studies (17,18), yet with different summary statistics. Therefore, we recalculated them to allow for direct comparison with our genetic overlap analysis.

#### Conjunctional false discovery rate

We used conjunctional FDR (conjFDR) analyses to investigate genetic overlap between AN and BMI and between AN and age at menarche. Detailed information about the conjFDR statistical framework can be found in previous publications (32,33), and the software can be accessed online at https://github.com/precimed/pleiofdr/. We performed the analysis in MATLAB R2017b, and used the standard recommendation of FDR < 0.05 for our conjFDR threshold. Notably, the major histocompatibility complex (MHC) region was excluded before running conjFDR analyses (chr6:25,119,106–33,854,733).

#### Gene mapping and functional annotation

We uploaded the FDR statistics from our genetic overlap analysis to the FUMA platform (available here: https://fuma.ctglab.nl/). FUMA aims to facilitate the identification of most likely causal variants from GWAS results, and allowed us to functionally map and annotate the candidate SNPs we had identified using positional, eQTL, and chromatin interaction mapping (34). A detailed description of FUMA settings can be found in the **Supplements**. Novelty was assessed against related studies (17,18,25,35–39).

#### Neandertal selective sweep score

We used the Neandertal selective sweep (NSS) score to assess possible recent selection at genomic loci associated with AN (R 4.2.2; www.r-project.org). The NSS score measures the relative abundance of ancestral *vs.* non-ancestral alleles in Neandertals and humans, through the alignment of primates, Neanderthal and human consensus sequences – with a negative NSS score indicative of plausible positive selection in humans (28,40). The score can be downloaded from the UCSC genome browser (http://genome.ucsc.edu, ntSssZScorePMVar track (S-scores)) (28,40). A detailed description of the analytical procedure can be found in the **Supplements.**

## Results

### Anorexia Nervosa genetically overlaps with Body Mass Index and Age at Menarche

We identified significant polygenic overlap between AN and BMI, and between AN and age at menarche. For both BMI and age at menarche, the proportion of non-null SNPs in AN increased with higher levels of these phenotypes, and *vice versa*. **Figure 1** illustrates the corresponding stratified Q-Q plots and shows an increasing enrichment as the statistical threshold becomes more stringent, supporting polygenic overlap between AN and BMI, and between AN and age at menarche.

**Figure 1:**
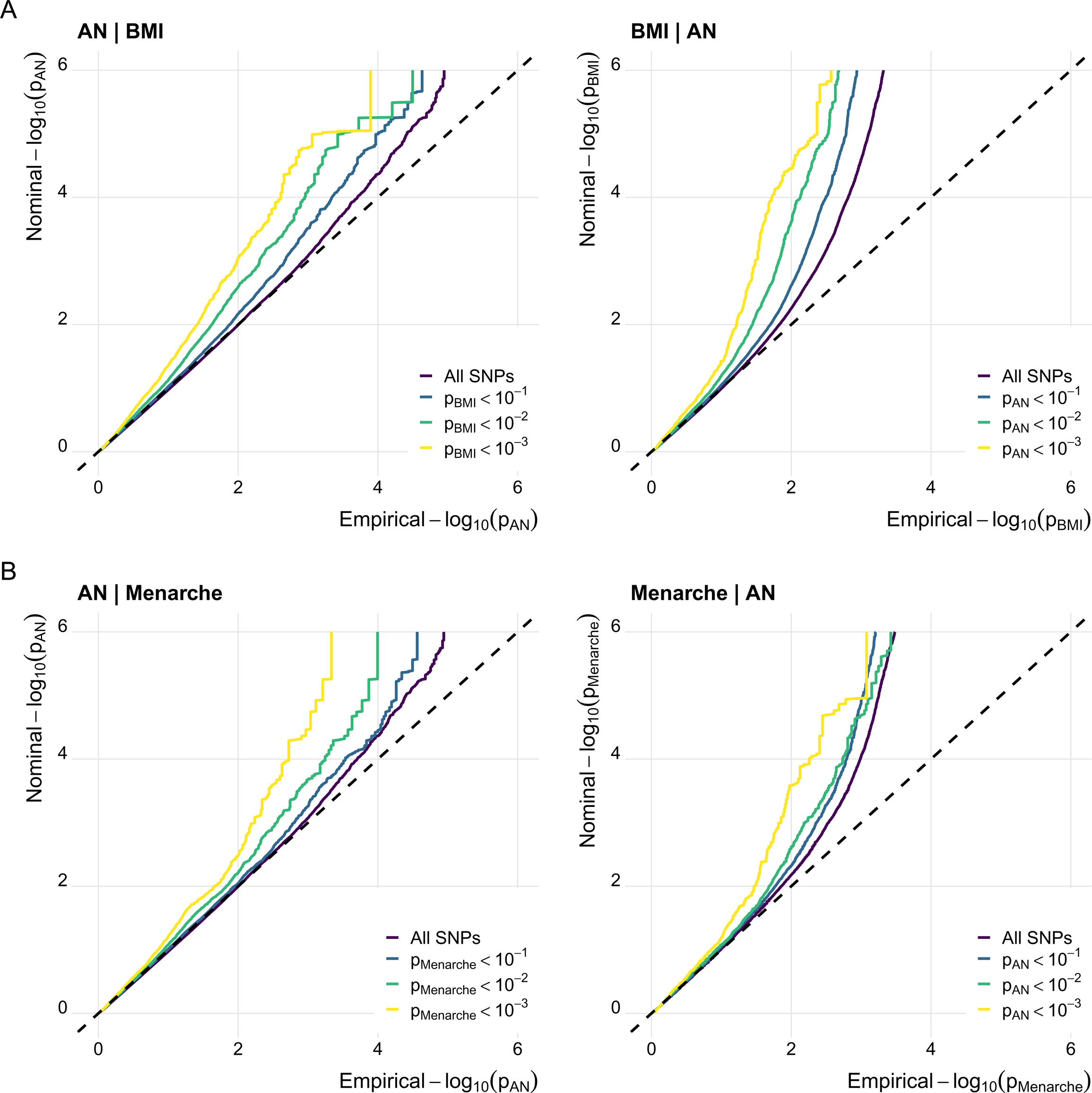
anorexia nervosa genetically overlaps with body mass index and age at menarche. <u>Figure legend</u>: Stratified quantile-quantile (QQ) plots displaying (A) increasing levels of SNP enrichment for AN conditioned on association *p*-values for BMI, and *vice versa*; (B) increasing levels of SNP enrichment for AN conditioned on association *p*-values for age at menarche, and *vice versa*

This overlap was further supported by statistically significant shared genomic loci between the phenotypes. Specifically, we identified 22 shared genomic risk loci between AN and BMI (67 independent significant SNPs), and nine shared genomic risk loci between AN and age at menarche (39 independent significant SNPs). Five loci overlapped between these two analyses, yielding 26 distinct loci overlapping with AN in total. Among the loci that overlapped between the two conjFDR analyses, three highlighted the same lead SNP or had corresponding independent SNPs (i.e., rs773949 (chr10), rs1351522 and rs34445652 (chr11) and rs141843711 (chr12)), possibly indicating that in these regions, the same variants may be relevant for the three traits.

To the best of our knowledge, seven of the 26 distinct risk loci have not previously been reported to be associated with AN. **Figure 2** show the corresponding Manhattan plot for the overlapping loci, while **Supplementary Tables 1-2** provide further details on each of them. For comparison, genetic correlation analysis did not capture the overlap between AN and age at menarche (r_g_=-0.03; *p* = 0.35), and only identified a link between AN and BMI (r_g_=-0.21; *p* = 7.3×10^-12^). These correlation estimates are consistent with a previous report (17) and illustrate genetic overlap in the absence of genetic correlation (see (41) for review).

**Figure 2:**
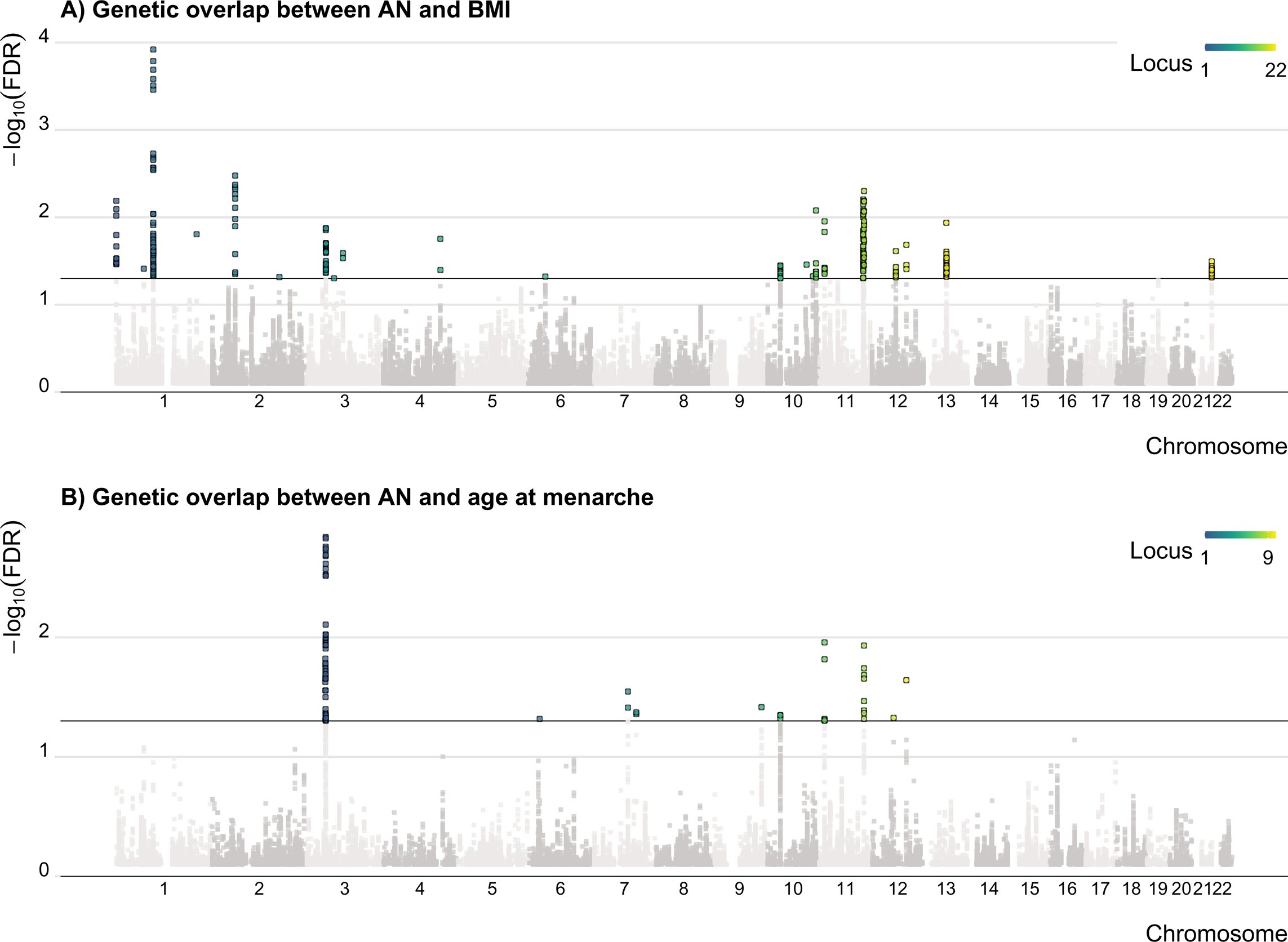
Identification of loci shared between anorexia nervosa and body mass index, and anorexia nervosa and age at menarche. <u>Figure legend:</u> Manhattan plots presenting in color the significant loci identified as shared between (A) AN | BMI and; (B) AN | Menarche.

### Specificity of the Genetic Overlap Between Anorexia Nervosa, Body Mass Index and Age at Menarche

A major challenge in psychiatry is the great heterogeneity and overlap across established diagnostic categories. Consequently, we verified whether the loci overlapping between AN, BMI and age at menarche were specific to AN, or more broadly associated with psychiatric disorders, by running additional analyses to assess the link between BMI or age at menarche and major depressive disorder (MDD) and schizophrenia. Detailed results are available as **Supplements**. In short, 59% of the loci shared between AN and BMI and 78% of the loci shared between AN and age at menarche could be considered as specific for AN, as they were not identified in the schizophrenia or MDD conjFDR analyses.

### Functional Annotation of Overlapping Loci between Anorexia Nervosa, Body Mass Index and Age at Menarche

The majority of the SNPs identified in the genetic overlap analyses were located in intronic and intergenic regions (**Supplementary Figure 5**). For both analyses, variants were mapped to genes with enriched expression in brain tissues and skeletal muscle (mostly downregulation; **Supplementary Figure 6-7**). Functions of these genes included neurodevelopmental, metabolic and cellular processes (**Supplementary Figures 8-11**).

One of the new loci that we identified was shared between AN and age at menarche (locus 5) and between AN and BMI (locus 12) and mapped to *PARD3*, which is involved in various neurodevelopmental, cellular, metabolic and developmental processes and has been linked to schizophrenia. We also identified *ARNTL*, known for its importance in the regulation of sleep, weight and eating behavior, as part of a locus shared between AN and age at menarche (locus 6) and between AN and BMI (locus 16).

In the analysis between AN and BMI, most of the genes were related to neurodevelopmental processes and overlapped with mental health-related phenotypes, such as mood swings, schizophrenia, neuroticism and risk-taking. Other mental health problems previously associated with these genes include bulimia nervosa and MDD (*NEGR1* (42,43)) autism spectrum disorders (*SLC25A12,* (44)) and suicide attempts (*ADARB1* (45)). Furthermore, some of these genes (e.g., *NEGR1, PTBP2, BCL11A, ARNTL, SLC25A12, ADARB1*) were identified in a recent study investigating the genetic overlap between AN and mental disorders (38), further supporting their relevance to AN.

Finally, the analysis between AN and age at menarche identified a new locus for AN that did not overlap with those identified in the analysis between AN and BMI. This locus was close to the *NR5A1* gene, which is involved in gonad development, hormonal mediated signalling pathways and aging.

### Possible Evolutionary Selection at Loci Associated With Anorexia Nervosa

Our analysis of Neanderthal Selective Sweep scores revealed that gene variants related to AN may have been under recent evolutionary drives in humans. The QQ-plot in **Figure 3** illustrates enrichment of AN-related SNPs for more negative NSS scores, where negative NSS scores point toward recent positive evolutionary selection.

**Figure 3:**
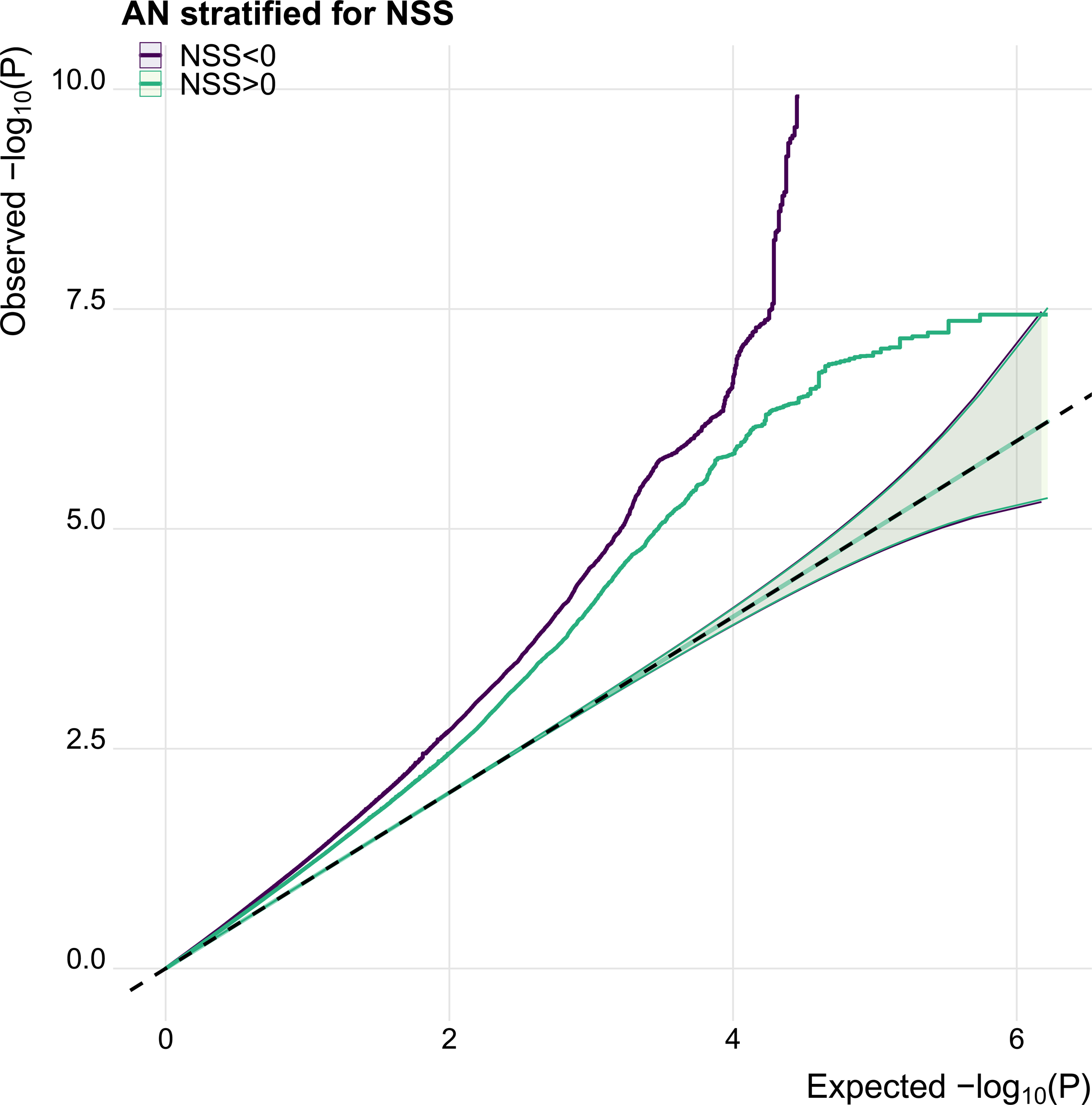
Comparison to Neanderthal genome suggests recent evolutionary selection for SNPs associated with anorexia nervosa. <u>Figure legend:</u> Quantile-Quantile (QQ) plot showing the enrichment of more negative Neanderthal Selective Sweep scores in SNPs associated with AN, pointing toward recent positive evolutionary selection.

We next assessed NSS score patterns for BMI and age at menarche, and for SNPs overlapping between these phenotypes and AN. NSS score values were available for 111 of the 275 candidate SNPs identified as overlapping between AN and BMI (40%), and for 38 of the 92 candidate SNPs overlapping between AN and age at menarche (41%). First, we compared the distribution of NSS scores for SNPs associated with AN, BMI and those identified as shared between AN and BMI (**Figure 4A**). We found that AN candidate SNPs were enriched for negative NSS scores (mean: -1.91, sd: 1.61) compared to BMI candidate SNPs (mean: -0.10, sd: 2.00), with candidate SNPs overlapping between AN and BMI standing in-between (mean: -0.62, sd: 1.73). We repeated this approach for AN and age at menarche and found similar patterns as above, that is more negative NSS scores for AN candidate SNPs compared to SNPs associated with age at menarche (mean: -0.63, sd: 1.71), and with SNPs overlapping between AN and age at menarche (mean: -1.44, sd: 1.94) standing in-between (**Figure 4B**). To put these results into perspective, we performed a conjFDR analysis to identify genetic overlap between BMI and age at menarche, merged our results with the NSS score data and compared the NSS score distributions for candidate SNPs associated with BMI, age at menarche or BMI and age at menarche. We found that the distribution of NSS scores between BMI and age at menarche was similar and less skewed to the left (i.e. less negative NSS values) than what we observed for candidate SNPs associated with AN (**Figure 4C**), further supporting that recent evolutionary selection may have played a particular role in AN-associated variants.

**Figure 4:**
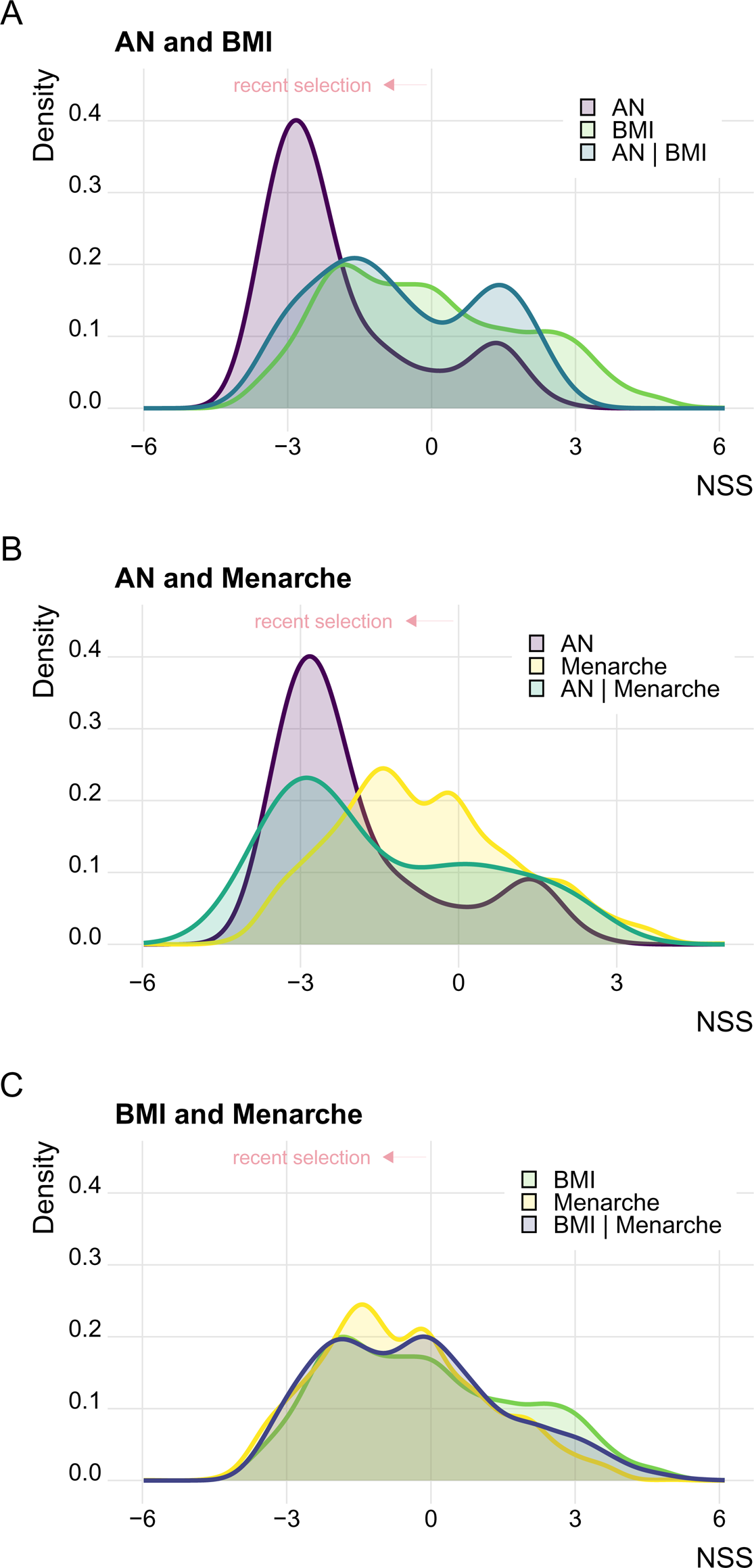
Compared to body mass index and age at menarche, SNPs associated with anorexia nervosa appear to be under stronger evolutionary selection. <u>Figure legend:</u> Distribution of Neanderthal Selective Sweep scores for candidate SNPs associated with (A) AN, BMI and those shared between AN | BMI; (B) AN, age at menarche and those shared between AN | Menarche; (C) BMI, age at menarche, and those shared between BMI | Menarche. Negative Neanderthal Selective Sweep scores point toward recent positive evolutionary selection.

### Evolutionary Placement of AN-associated Loci and Genes

Of all the identified loci, we investigated whether some were more likely to be subject to recent evolutionary dynamics. At the locus level, we found that four AN-associated loci overlapped with regions with the 5% smallest NSS mean score across the genome. These include the locus that mapped to the *PTBP2* gene (chromosome 1), two loci on chromosome 3 (one multigenic locus including *DAG1*, *RBM6*, *CCDC36*, *NCKIPSD*, *USP4* and *USP19*; and one intergenic locus in the vicinity of *NSUN3*) and one locus on chromosome 12. Two of these loci (on chromosome 3 and chromosome 12) also overlap with three regions identified by another evolutionary methods (i.e., Composite of Multiple Signal) as potential candidates for evolutionary adaptation (46), further supporting that these regions may have been involved in recent human evolution. Finally, we looked for overlap between AN-associated loci and previously identified differently methylated regions between humans and Neanderthals (47). We found that two AN-associated loci overlapped with such regions (including the multigenic loci on chromosome 3 mentioned above). This result supports that these regions possibly played a role in the adaptation of modern humans to their environment. More details on these loci can be found in **Supplementary Table 7**.

We next ranked genes by their evolutionary signal by calculating mean NSS scores per gene, as shown in **Figure 5**. For the main AN-GWAS, the genes with the most negative NSS scores were *C3orf84* and *CCDC71*. *CCDC71* is involved in lipid metabolism and fat cell differentiation, and according to our FUMA results, has been linked to autoimmune disorders of the gastrointestinal system. In the analysis of genetic overlap between AN and BMI, the gene with most negative NSS score was *GNB1* (chromosome 1), which is involved in cell-cell signaling. Mutations in *GNB1* have been linked to growth and (neuro)developmental delay, and gastrointestinal problems (48). In the overlap between AN and age at menarche, *USP4* (chromosome 3) had the most negative NSS score.

**Figure 5:**
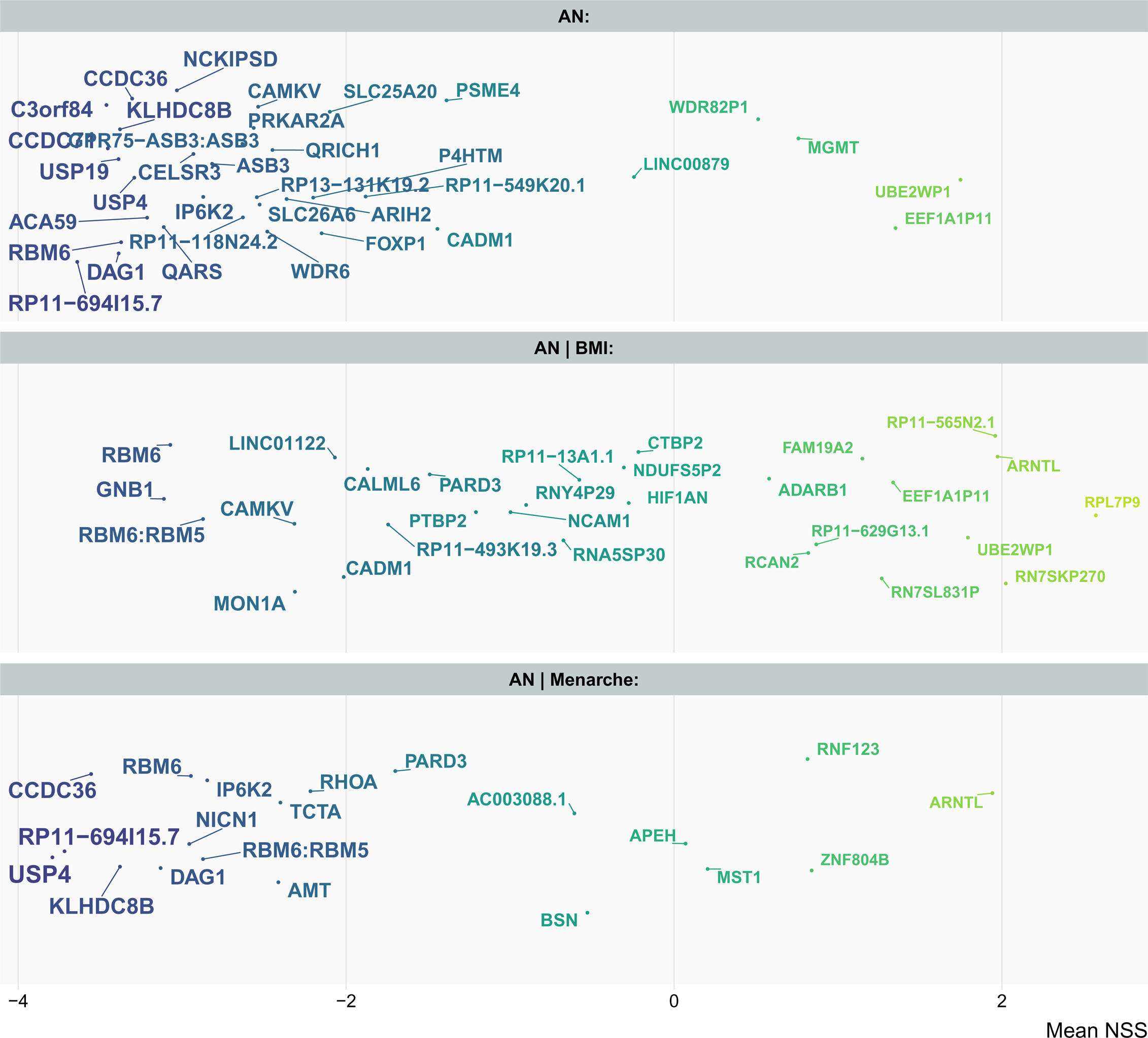
Genes associated with anorexia nervosa, highlighted from an evolutionary perspective. <u>Figure Legend:</u> Genes mapped to the loci associated with AN (top panel), AN | BMI (middle panel) or AN | Menarche (bottom panel). The x-axis represents Neanderthal Selective Sweep scores, with more negative Neanderthal Selective Sweep scores pointing toward recent positive evolutionary selection. Neanderthal Selective Sweep score per gene was determined as an average of the SNPs from which the gene was mapped. Location on y-axis is random (jitter) and was used for legibility of otherwise overlapping text. Darker colours are used to highlight genes with the lowest mean NSS scores.

A common pattern that emerged among many genes with strongly negative NSS was the implication of chronotype as an associated process. For example, *GRP7-ASB3* has been linked to chronotype and excessive daytime sleepiness, while *CAMKV*, *IP6K2* and *RHOA* have been associated with morning-ness, and *MONIA, RHOA, TCTA, AMT, NICN1, CAMKV* relate to sleep duration.

## Discussion

The present study established polygenic overlap between AN, BMI and age at menarche and through its overlap identified seven novel loci associated with AN. Further, we found that the genetic risk loci associated with AN may have been subject to recent evolutionary selection. The current findings deepen our knowledge of the genetics of AN and may help to better understand the biological mechanisms involved in this debilitating disorder.

Our analysis of genetic overlap between AN and BMI identified 22 significantly overlapping risk loci. Previous research has already established the inverse genetic correlation between AN and BMI, and that fat mass shows one of the strongest genetic correlations with AN (17). These correlations and the here identified overlap support that energy balance may be impaired in people with AN. Yet, the association between AN and BMI is likely not solely linked to energy balance, as a large proportion of the genes identified as overlapping between AN and BMI are relevant to brain functions. Among these genes, the identification of *NEGR1* as overlapping between AN and BMI (locus 2) is noteworthy, as *NEGR1* is involved in neurodevelopment and has been associated with obesity (30), age at menarche (49) and other psychiatric disorders (42,43). Recently, a variant in this locus was identified as shared between AN and MDD, and between AN and schizophrenia (38). An epigenome-wide study also identified differently methylated regions between people with active AN compared to people in remission from AN or healthy controls at the *NEGR1* gene locus (50), providing further support for its relevance for AN. Overall, our findings reinforce that studying EDs together with metabolic conditions like overweight/obesity would be beneficial, as their underlying (neuro)biology may overlap (51).

Puberty is an important time period for the development of EDs and may underline some of the sex differences in their prevalence and clinical presentation (52). Here, we identified nine significant loci overlapping between AN and age at menarche. This overlap is noteworthy given their weak, non-significant genetic correlation (17). These results may indicate that some variants may have an opposite effect on AN and age at menarche, or that different subtypes of AN (e.g. restrictive *vs.* binge/purge or early *vs.* late onset) may be differently associated with age at menarche. In this regard, a previous study found that an earlier age at menarche was linked to an earlier age at onset of AN in people with early-onset AN, but found no association between age at menarche and the risk of AN or with late-onset AN (25). These findings may be somewhat surprising given previous evidence of delayed growth and sexual maturation in youths with AN (23,24). Yet, this may also indicate that different AN subtypes may have diverse biological underpinnings, especially given that the literature on AN and age at menarche remains scarce, with few authors considering AN subtypes, and sample sizes being often limited. Further research including puberty-related variables as a primary focus (and not just as confounders) is therefore warranted.

A new finding from the present study is that several genes overlapping between AN, BMI and age at menarche have in common that they are related to circadian-related phenotypes. This includes *ARNTL*, a major component of the circadian clock system (53,54). In animal models, knock-out or reduced expression of *ARNTL* have been associated with altered circadian rhythms, body weight, alterations in locomotor activity and eating behaviours, delayed puberty and infertility, cognitive deficits and shortened lifespan (54–56). Given their role in energy homeostasis and their mutual exclusivity, sleep patterns and eating behaviors are closely related; however, studies on the clinical association between AN and sleep disorders are scarce. Existing evidence suggests that sleep problems, including insomnia, are common in people with EDs (57–60) and that sleep disruptions can worsen the severity of AN (60,61). Poor sleep and altered circadian rhythms have also been linked to metabolic dysregulation and obesity (62,63), and to woman’s reproductive health (55,56). One might therefore wonder whether the circadian cycle might be a key element linking AN, BMI and puberty, and whether this phenotype might have conferred some sort of evolutionary advantage. Depending on environmental conditions, being more or less “restless” might have impacted the risk to suffer from malnutrition and might have affected the chances of reproductive success. More studies are needed to clarify this, but given that AN remains excessively difficult to treat, and considering the results of the present study and of other recent works (64,65), the clinical link between AN, chronobiology and sleep, as well as a the biology behind it should be explored as a potential starting point for improving interventions.

Studying AN from an evolutionary perspective revealed that some of the genomic regions associated with AN might have been involved in human evolution, as suggested by an enrichment in negative NSS scores. Although more studies are needed to confirm this finding, it would align with studies in other fields that have suggested various metabolic adaptations after exposure to developmental adversity. For example, longitudinal studies on the Dutch Famine and Chinese Hunger cohorts have shown that individuals who were exposed to hunger during pregnancy had a higher risk of long-term psychiatric and metabolic disorders (66–68). Some have hypothesised that these effects could be mediated by epigenetic mechanisms (69). Overall, some genetic or epigenetic adaptations conferring a metabolic or reproductive advantage in a given environment may now also increase the risk of certain disorders in today’s living conditions, including AN.

The present study has some important strengths, including the use of statistical methods allowing to explore genetic pleiotropy and the use of Neanderthal genomic data to employ an evolutionary lens on AN. Yet, some limitations should be considered. Merging the GWAS data with the NSS data left us with several SNPs for which NSS scores were not available and thus some AN-associated signal could not be explored in terms of evolution. Furthermore, most retained SNPs had a negative NSS score (pointing toward recent selection), and we cannot exclude that the enrichment of negative NSS scores at SNPs related to AN may have other explanations. For example, some of the variants may not confer an evolutionary advantage but may instead have been inherited along with other variants with beneficial functions, through linkage disequilibrium. We circumvented this potential issue by comparing NSS distributions between phenotypes, which overall suggests more pronounced effects for AN. Further, four of the AN-associated loci overlapped with regions of the genome with the 5% smallest NSS scores (28), and some of those regions have also been identified by others as being under plausible evolutionary drive (46,47), which is reassuring. Another limitation is that the current data did not allow us to study subtypes of AN or sex, although there may be relevant differences between these groups (70,71). Given the great heterogeneity in ED diagnostic categories, including within the AN diagnosis itself, an important question that remains is whether some of the associations highlighted here are specific to certain AN subtypes, or if they could be extended to other type of EDs. Phenotypically, there is evidence that sleep patterns may vary between ED categories (e.g., restrictive *vs.* purging (72)). Future genomic efforts in the field of EDs will eventually allow for a better understanding of the genetic subtilities underlying different categories, not only in regard to AN-R vs. AN-BP, but also other type of EDs. Finally, an important perspective for future work is the need for longitudinal studies, together with the use of other statistical frameworks to investigate causality in genetic associations between metabolism, sexual maturation and mental health (e.g., (73)). Ultimately, this will provide a better understanding of the developmental course and potential causal relationships between AN, BMI and age at menarche that could not be addressed in the present study.

In conclusion, the present study has identified genes that overlap between AN, BMI and age at menarche, and found that some of these genes may have been undergoing recent evolutionary selection. We identified genes relevant to circadian rhythms in the association signal between AN, BMI and age at menarche, and with Neanderthal scores pointing at recent selection. Our study emphasizes the utility of employing an evolutionary lens in psychiatric genetics and the results identify candidates for future studies, such as detailed investigations into the genetic link between chronobiology and AN. As such, analysis of pleiotropy and evolutionary context may go hand in hand in further deciphering the mechanisms underlying this debilitating disorder.

## Data Availability

All data are available online from the respective consortia

https://www.reprogen.org/data_download.html

https://portals.broadinstitute.org/collaboration/giant/index.php/GIANT_consortium_data_files

https://pgc.unc.edu/for-researchers/download-results/

## Acknowledgements

This work was undertaken on the Tjeneste for Sensitive Data (TSD) facilities which is operated/developed by University of Oslo TSD service group (IT-Department), and owned by the University of Oslo as well as on resources provided by the National Infrastructure for High Performance Computing and Data Storage in Norway (UNINETT Sigma2). We would like to thank the three consortia (PGC, GIANT and ReproGen) from which we obtained summary statistics, and their respective research participants, for making this work possible. The present manuscript was published as a preprint on medRxiv (doi.org/10.1101/2023.08.09.23293879).

