## Supplements for "An Evolutionary Perspective on the Genetics of Anorexia Nervosa"

Table of Content

### Supplementary Methods

#### Sample

To avoid sample overlap, we selected GWAS excluding participants from the UK Biobank for the BMI and age at menarche phenotypes. The AN GWAS was conducted by the Psychiatric Genomics Consortium and covered data from n = 16,992 individuals with AN and n = 55,525 healthy controls. The BMI GWAS was conducted by the GIANT consortium using data from n = 339,224 individuals. For age at menarche, we accessed a GWAS from the ReproGen Consortium performed in n = 182,416 individuals.

#### Gene mapping and functional annotation – FUMA settings

The FUMA SNP2GENE function annotates SNPs with functional categories, CADD scores (deleterious protein effect indicated by a CADD > 12.37 (1)), RegulomeDB scores (probable regulatory functions indicated by a score < 1F (2)) and chromatin states (more open chromatin state, indicated by a score between 1-7, suggest a region is more accessible for epigenetic regulatory mechanisms (3)). In the present study, locus numbers reflect their positional order from chromosome 1-22. Further, we used the GENE2FUNC function to identify enriched gene-sets and overrepresented biological pathways. We considered SNPs with a conjFDR > 0.05 and a linkage disequilibrium (LD) R2 < 0.6 with each other as independent significant SNPs. Among these, SNPs with a R2 < 0.1 with each other were considered as lead SNPs. Candidate SNPs with a LD R2 ≥ 0.6 with the lead variants and with a conjFDR < 0.1 were submitted to FUMA.

#### Neandertal selective sweep score analyses

First, we used plink to clump candidate SNPs for the AN, BMI and age at menarche main GWAS (<https://github.com/precimed/python_convert>). Then, we extracted NSS scores for all candidate SNPs identified in the AN, BMI and age at menarche GWAS, as well as for all candidate SNPs identified through our analysis of genetic overlap between them. This allowed us to examine the likelihood of recent selection (i.e., more negative NSS scores are associated with increased likelihood of positive selection) for the candidate SNPs associated with AN. Quantile-Quantile (QQ) plots were created to investigate the enrichment of negative NSS scores in SNPs associated with AN. We also graphically explored the distribution of NSS scores for candidate SNPs identified in previous AN, BMI and age at menarche GWAS, as well as candidate SNPs overlapping between the traits.

In addition to looking for enrichment in negative NSS scores in SNPs associated with AN, BMI or age at menarche, we assessed the overlap between the genomic loci associated with AN and the genomic regions identified by Green & al. (4) as having the 5% smallest NSS score (i.e., regions with the strongest signal for potential positive selection, corresponding to a NSS score ≤ -4.32). These regions are available from UCSC genome browser (http://genome.ucsc.edu, 5% Lowest S Track).

Lastly, to validate our results, we tested whether there was overlap between AN-associated loci and regions previously identified by two other evolutionary methods. The first of these methods is the Composite of Multiple Signal (CMS) (5). Of note, we converted genomic coordinates using the LiftOver tool (available at: <https://genome.ucsc.edu/cgi-bin/hgLiftOver>) to match the CMS regions identified as having strong signal for selection with the AN-associated loci. The second method uses a different angle to support the importance of specific regions in the adaptation of humans to their environment, as it is based on epigenetics. The author of this study identified genomic loci marked by methylation differences between modern humans and Neanderthals (6), and we assessed the overlap between these regions and the AN-associated loci

### Supplementary Results

#### Genetic overlap between body mass index and age at menarche and other psychiatric disorders

To investigate for the here identified overlap between BMI/age at menarche and AN to what degree these variants are specific to AN, we drew comparison to other disorders. We ran additional conjFDR analyses to assess the link between BMI or age at menarche and major depressive disorder (MDD) and schizophrenia, using publicly available GWAS summary statistic for those traits (7,8). We chose MDD and schizophrenia because these disorders have among the best powered GWAS in psychiatry to date. Given known associations between BMI, puberty and mental health, we expected to find genetic associations with other psychiatric traits. We hypothesised that using more powerful GWAS might allow us to find a substantial amount of associations with BMI and age at menarche; and thus, provide a good way to assess the specificity of our findings for AN. As shown in **supplementary figures 1 and 2**, the stratified Q-Q plots suggest an enrichment in significant associations between the four combinations of traits. We identified 104 significantly overlapping loci between MDD and BMI, 147 significantly overlapping loci between schizophrenia and BMI, 54 significantly overlapping loci between MDD and age at menarche, and 73 significantly overlapping loci between schizophrenia and age at menarche (**Supplementary figures 3 and 4, Supplementary tables 1-6)**. Looking at the overlap between these loci and those found to overlap between AN and BMI, 13 of 22 loci (59%) shared between AN and BMI could be considered specific to AN, as they were not identified in the schizophrenia | BMI or MDD | BMI analyses (**Supplementary table 1**). For age at menarche, two of the nine loci that we identified for AN were also found for MDD or schizophrenia, indicating in turn that 78% of our hits could be considered as specific to AN (**Supplementary table 2**).

### Supplementary Figures

##### Supplementary Figure 1: major depressive disorder genetically overlaps with body mass index and age at menarche

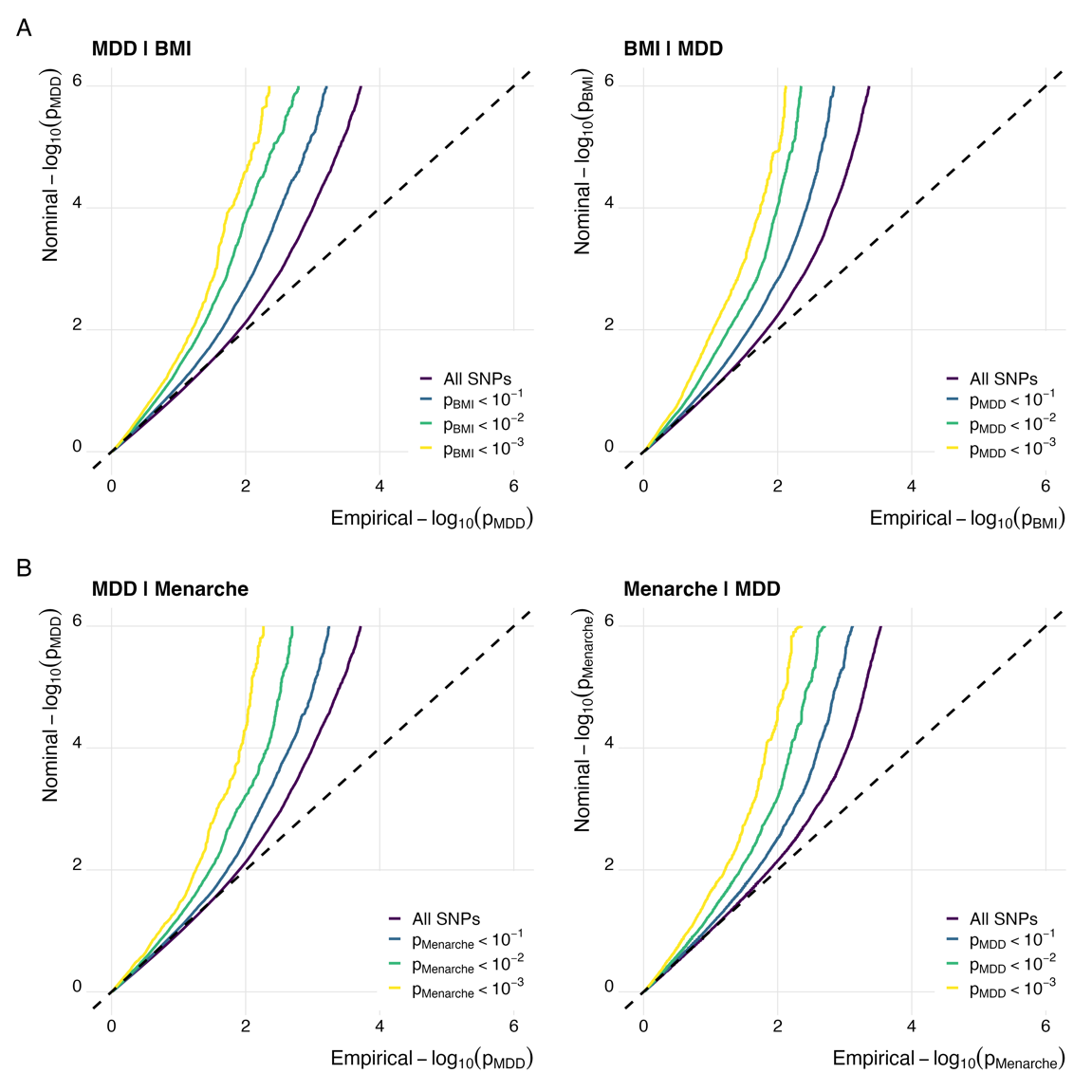

Figure legend: Stratified quantile-quantile (QQ) plots displaying (A) increasing levels of SNP enrichment for MDD conditioned on association p-values for BMI, and vice versa; (B) increasing levels of SNP enrichment for MDD conditioned on association p-values for age at menarche, and vice versa. SNP, singe nucleotide polymorphism; MDD, major depressive disorder; BMI, body mass index.

##### Supplementary Figure 2: schizophrenia genetically overlaps with body mass index and age at menarche

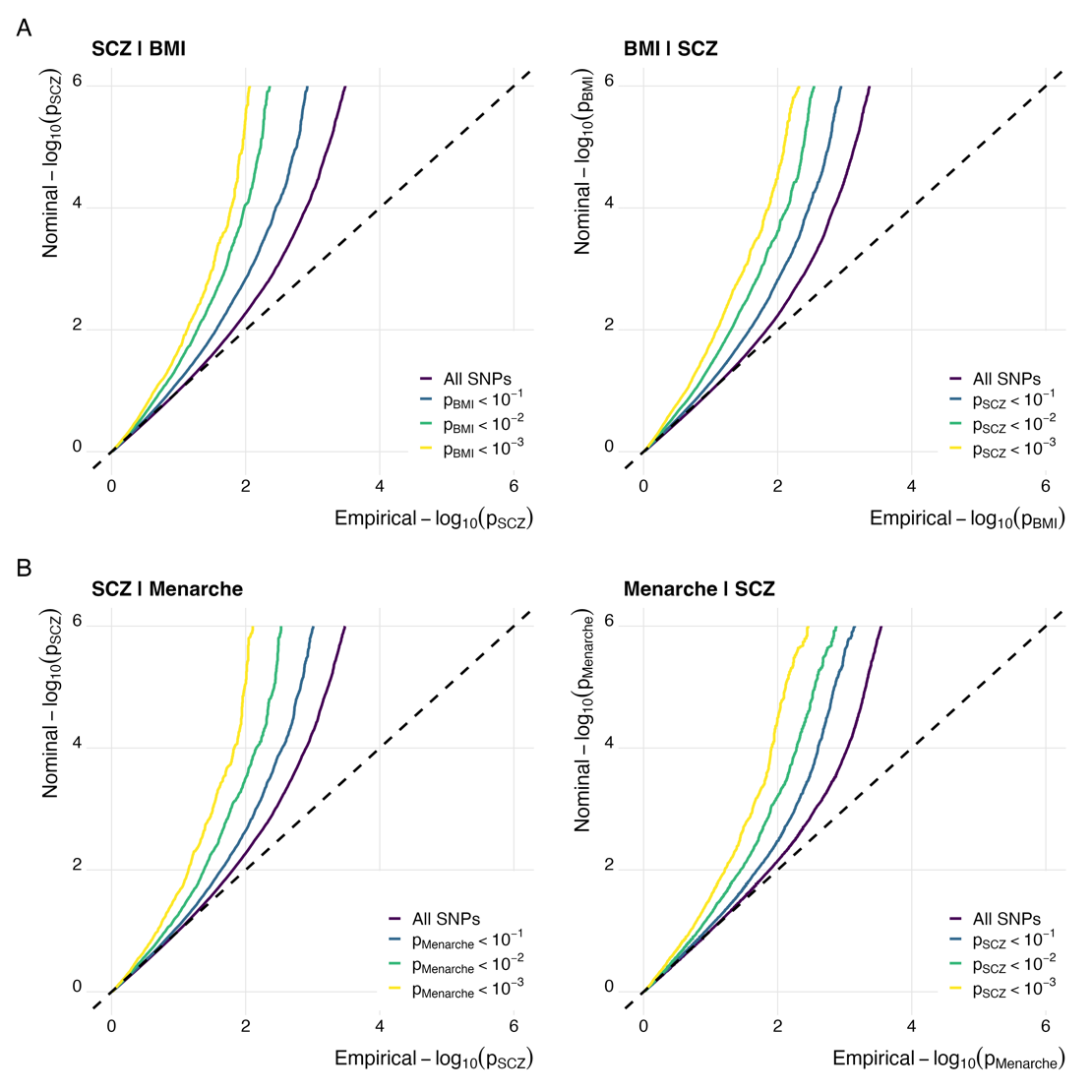

Figure legend: Stratified quantile-quantile (QQ) plots displaying (A) increasing levels of SNP enrichment for SCZ conditioned on association *p*-values for BMI, and *vice versa*; (B) increasing levels of SNP enrichment for SCZ conditioned on association *p*-values for age at menarche, and *vice versa*. SNP, singe nucleotide polymorphism; SCZ, schizophrenia; BMI, body mass index.

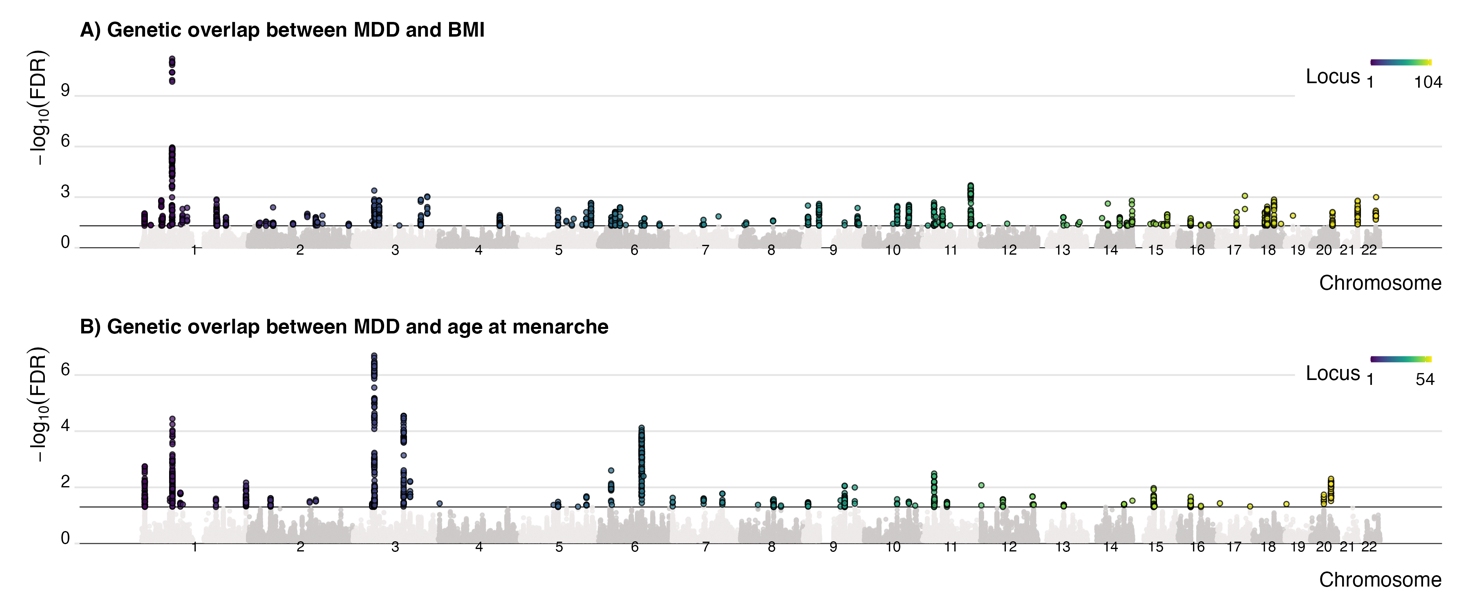

##### Supplementary Figure 3: identification of loci shared between major depressive disorders and body mass index, and major depressive disorder and age at menarche

Figure legend: Manhattan plots presenting in color the significant loci identified as shared between (A) MDD | BMI and; (B) MDD | Menarche. MDD, major depressive disorder; BMI, body mass index.

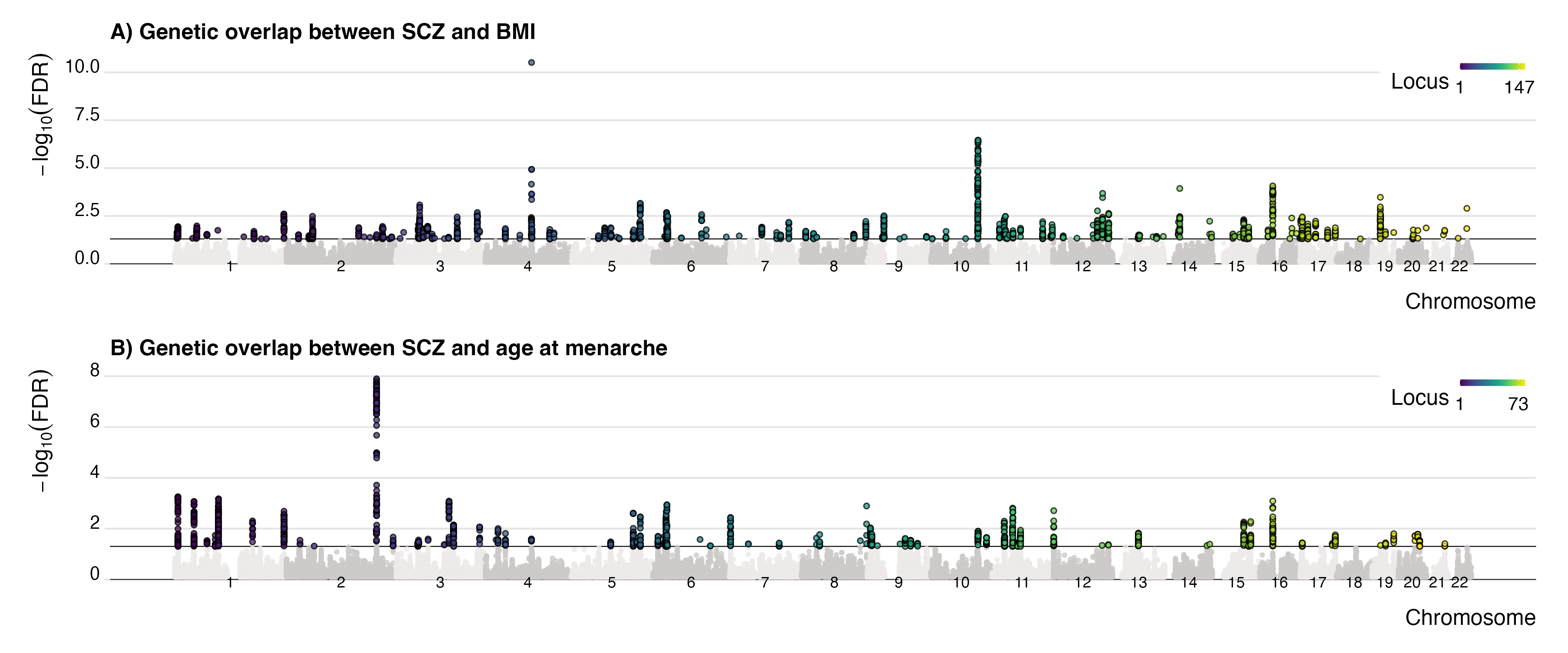

##### Supplementary Figure 4: identification of loci shared between schizophrenia and body mass index, and schizophrenia and age at menarche

Figure legend: Manhattan plots presenting in color the significant loci identified as shared between (A) SCZ | BMI and; (B) SCZ | Menarche. SCZ, schizophrenia; BMI, body mass index.

##### Supplementary Figure 5: Functional annotation of SNPs (A) shared between AN and BMI and; (B) shared between AN and age at menarche

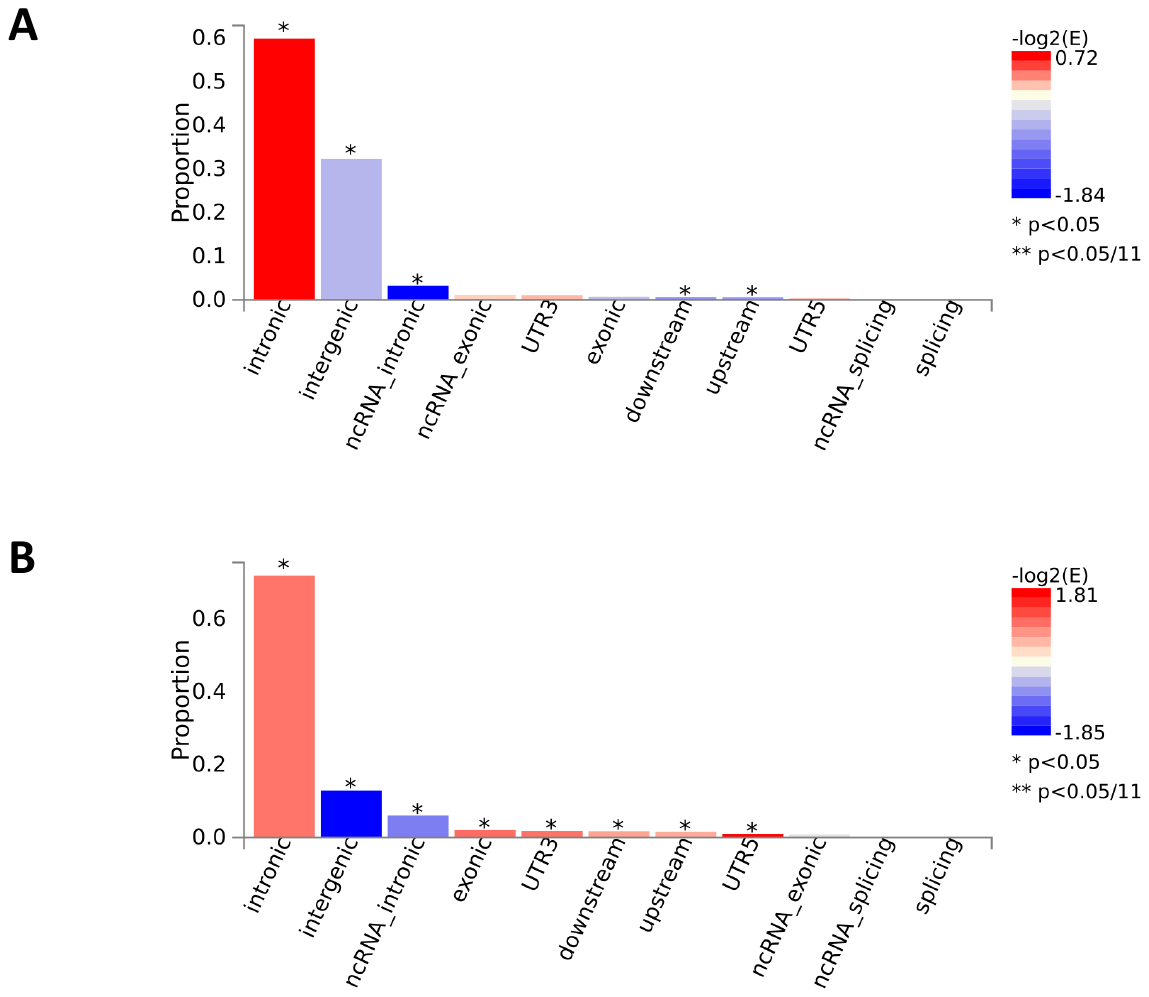

##### Supplementary Figure 6: Expression of mapped genes shared between AN and BMI

##### based on 54 tissue types

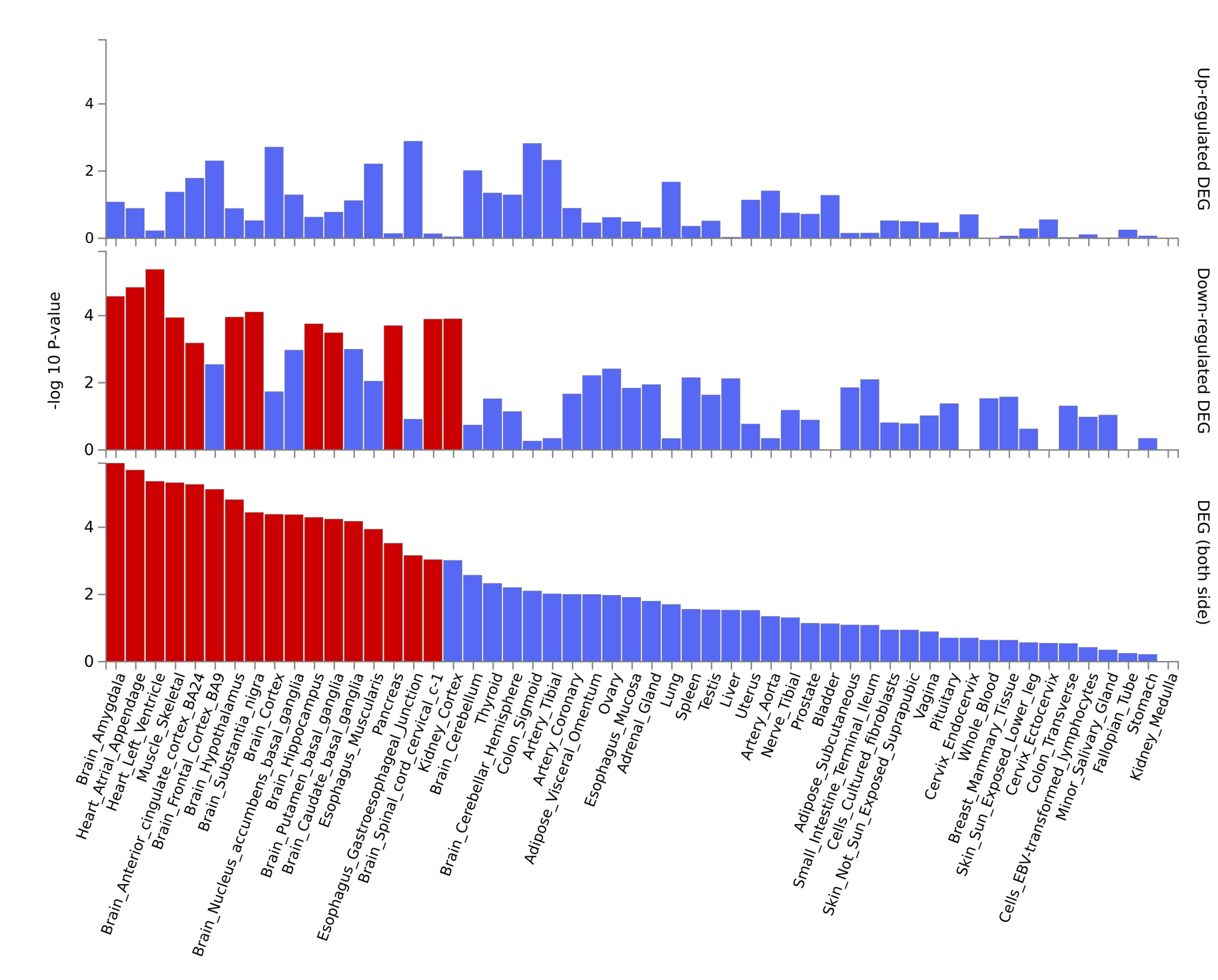

##### Supplementary Figure 7: Expression of mapped genes shared between AN and age at menarche based on 54 tissue types

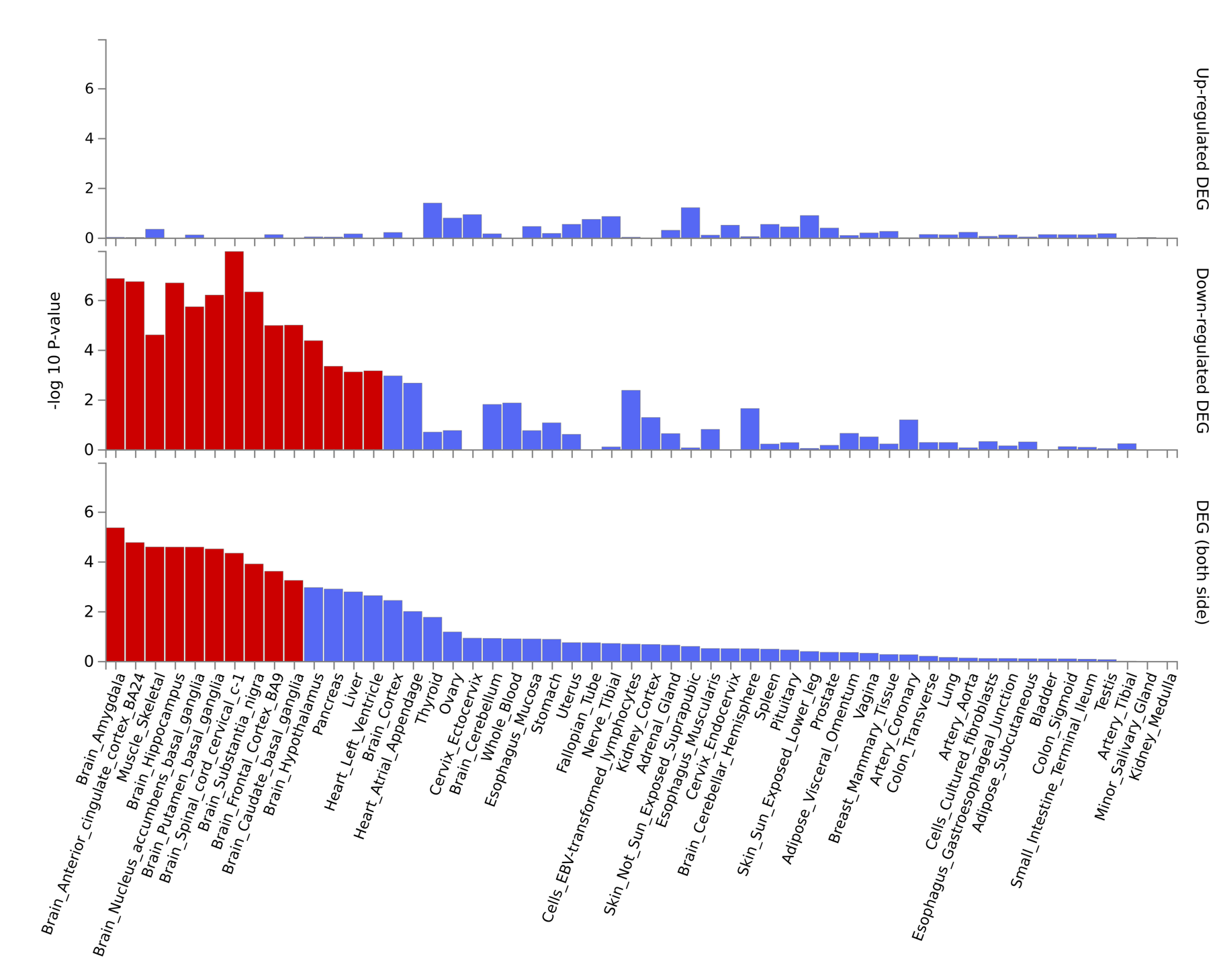

##### Supplementary Figure 8: GO biological processes identified with FUMA for the genes shared between AN and BMI

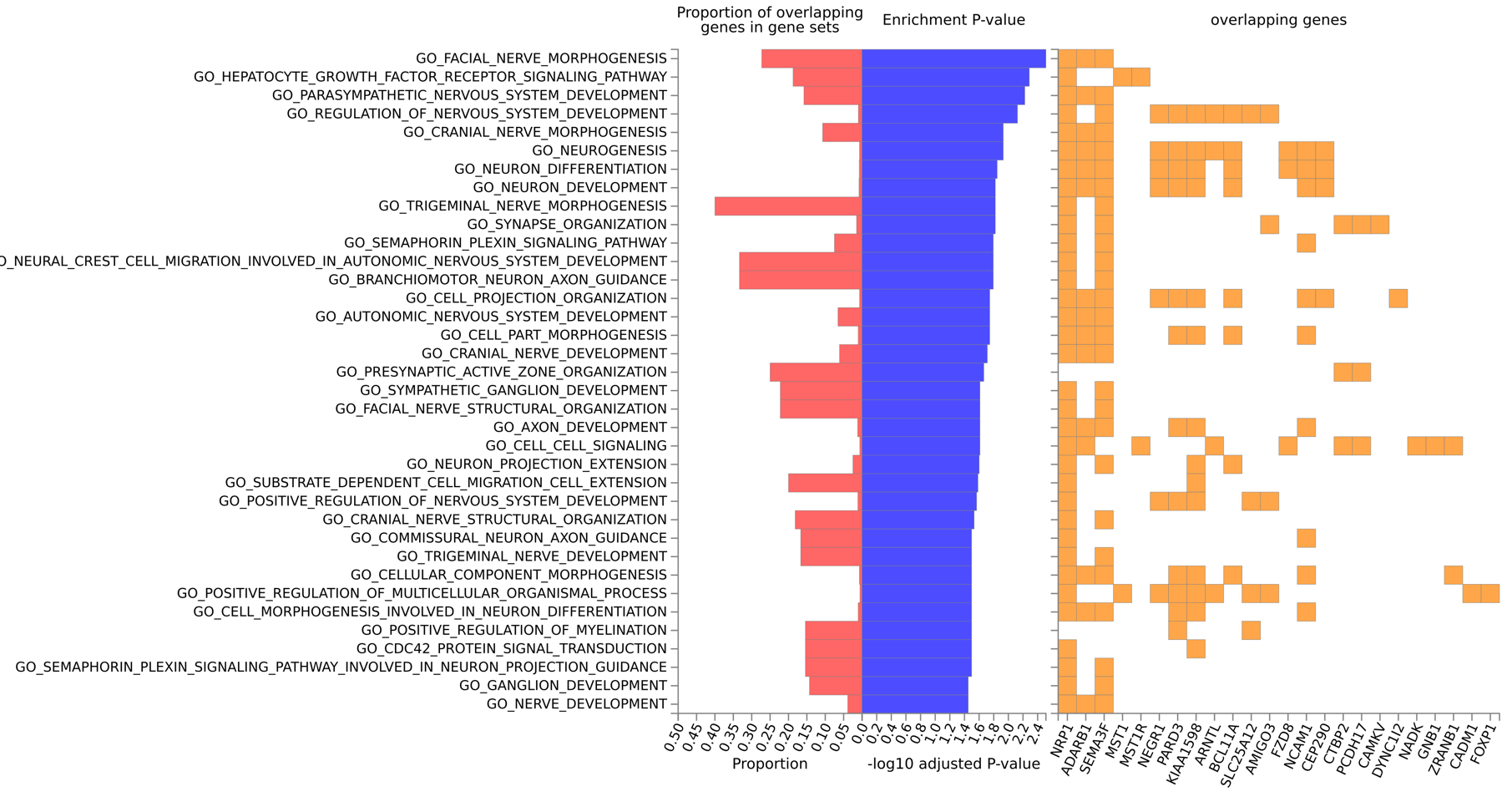

##### Supplementary Figure 9: GO biological processes identified with FUMA for the genes shared between AN and age at menarche

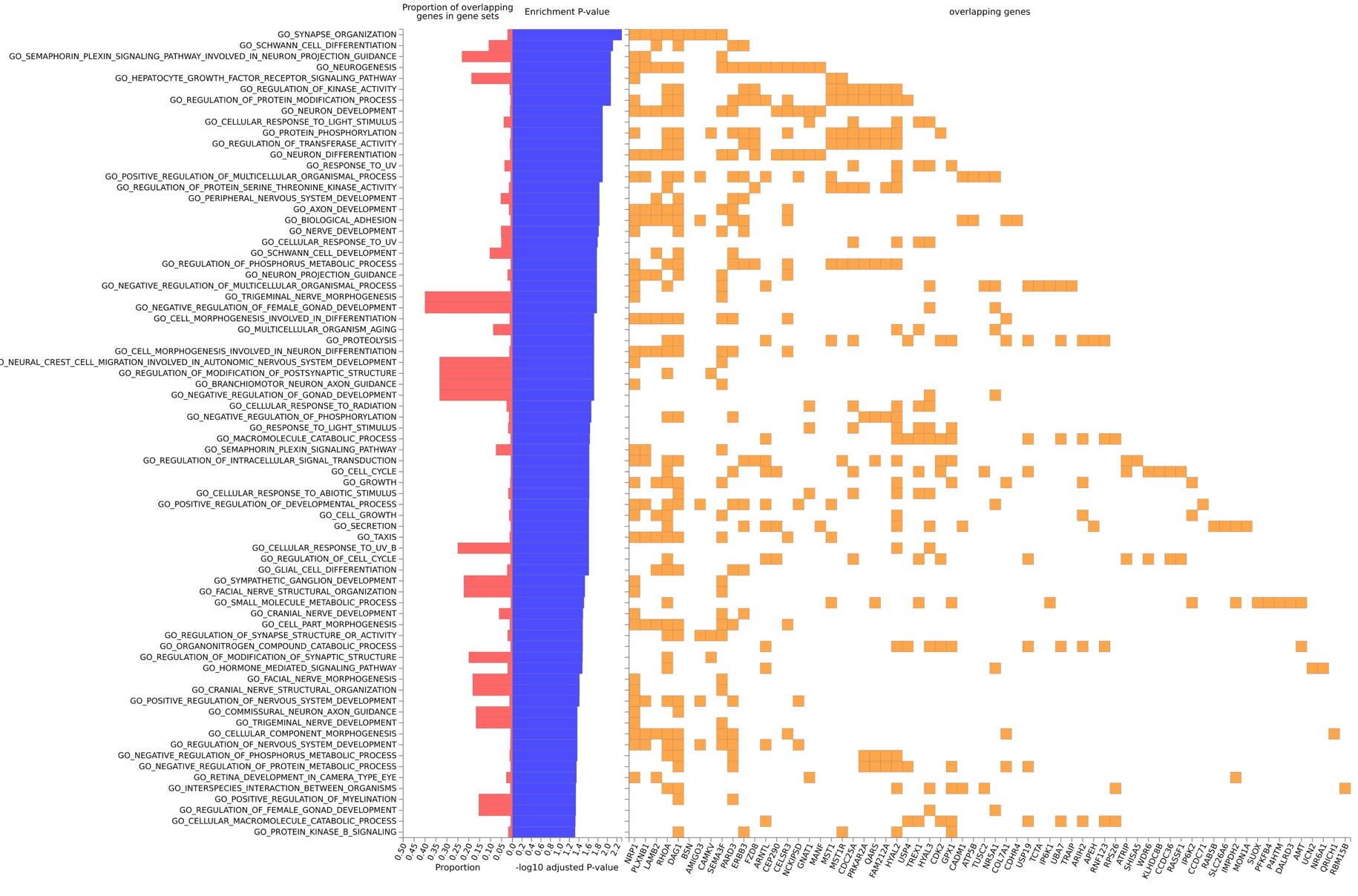

##### Supplementary Figure 10: GWAS phenotypes identified with FUMA for the genes shared between AN and BMI

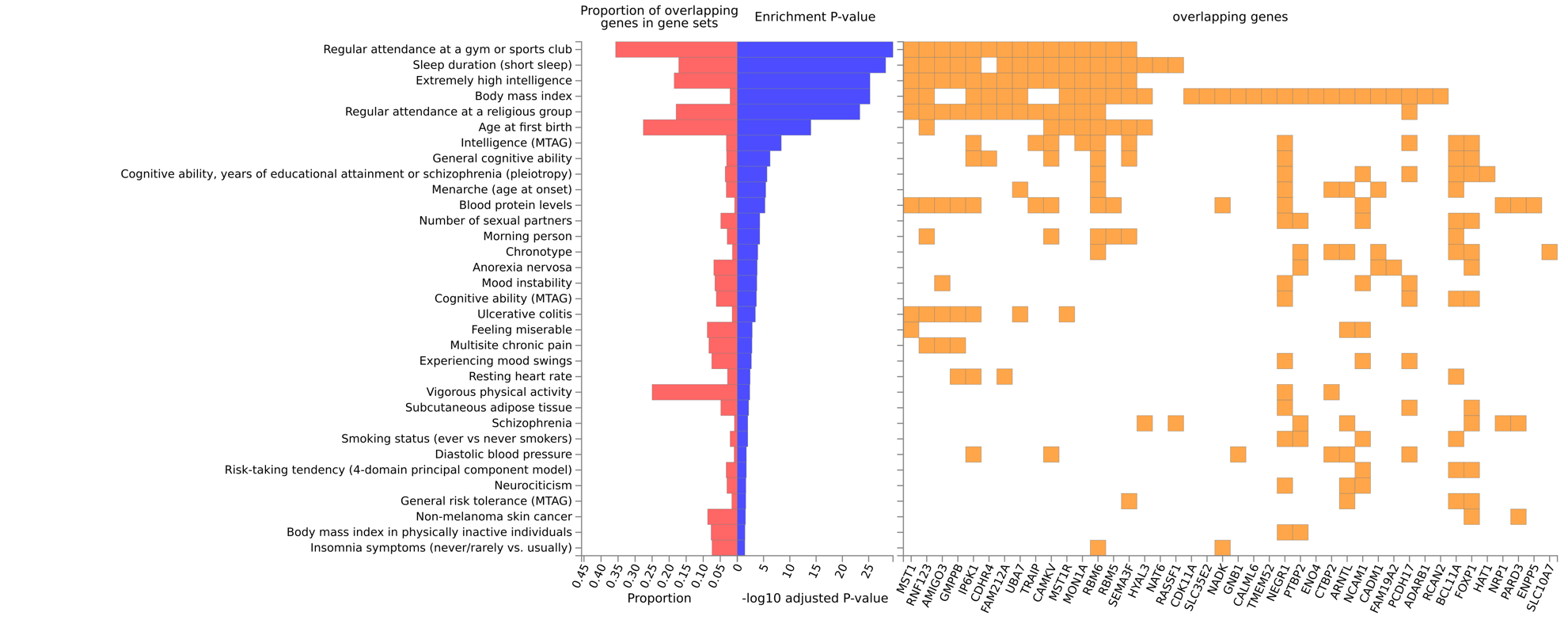

##### Supplementary Figure 11: GWAS phenotypes identified with FUMA for the genes shared between AN and age at menarche

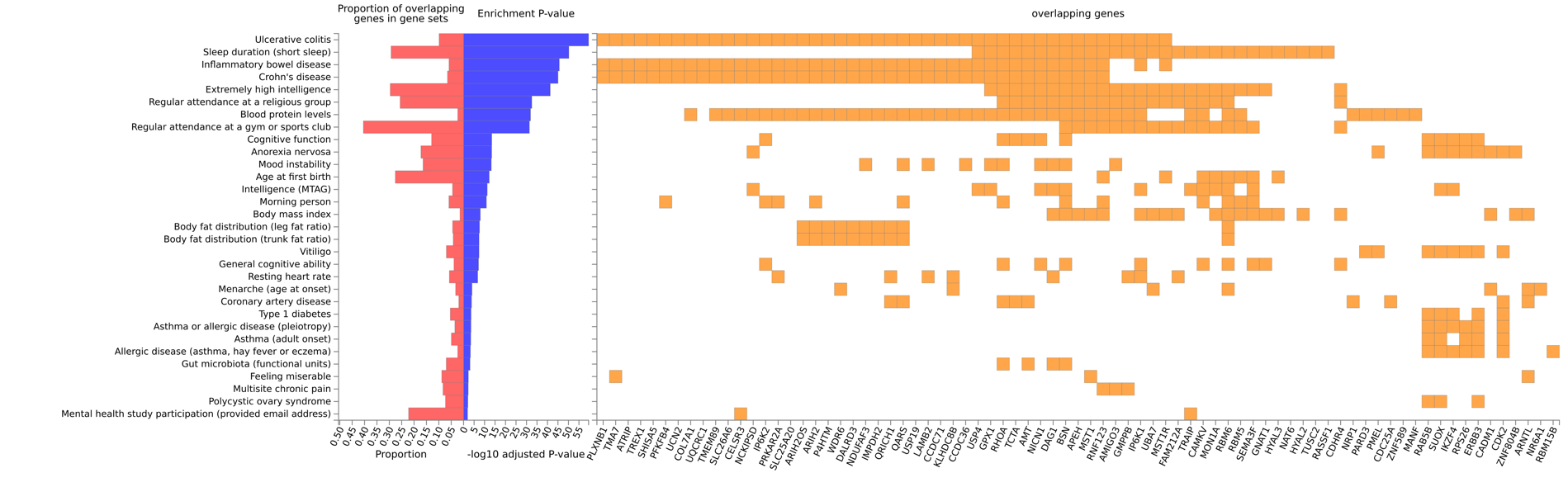

### Supplementary Tables

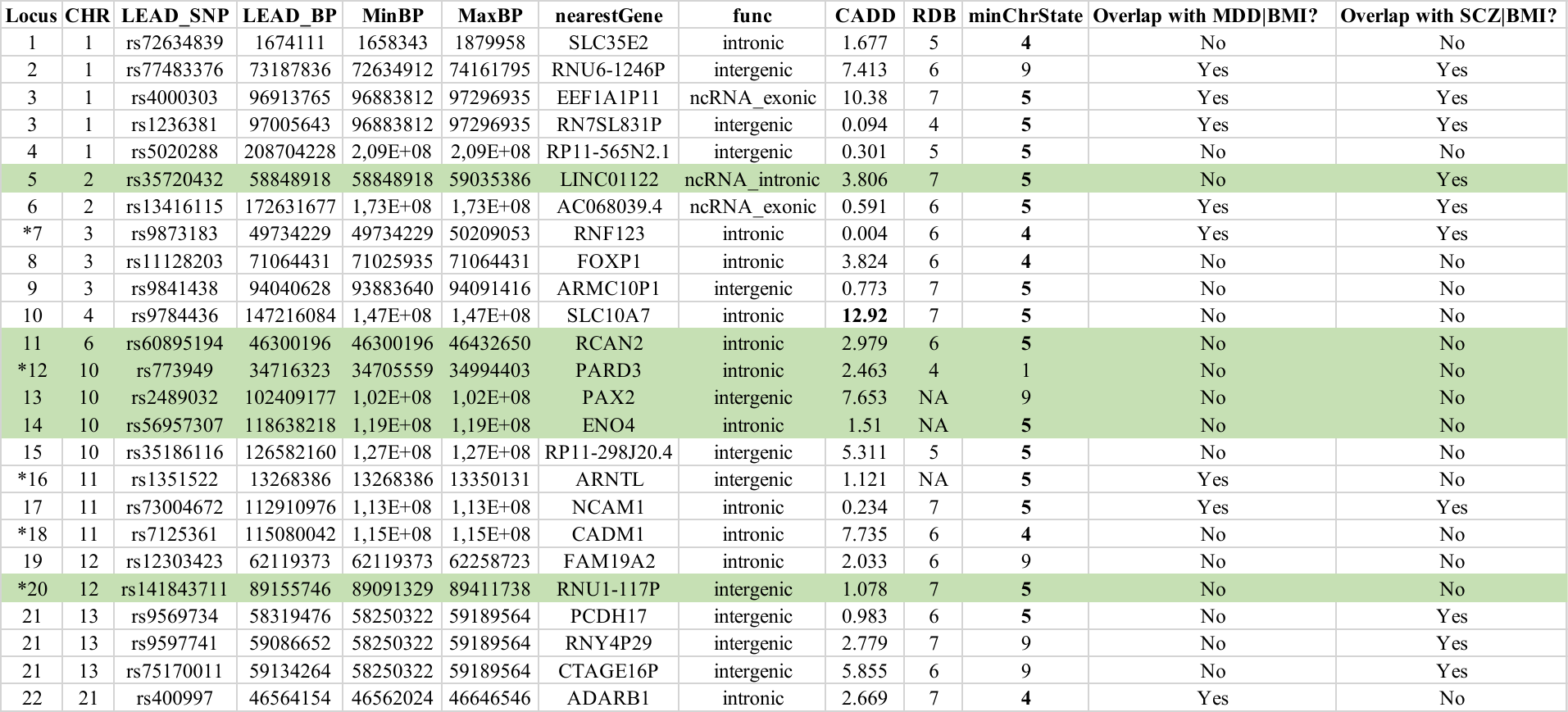

##### Supplementary Table 1: Risk loci shared between anorexia nervosa and body mass index

Loci not previously identified as associated with Anorexia Nervosa

*loci overlapping with those identified in the conjFDR analysis of AN and age at menarche

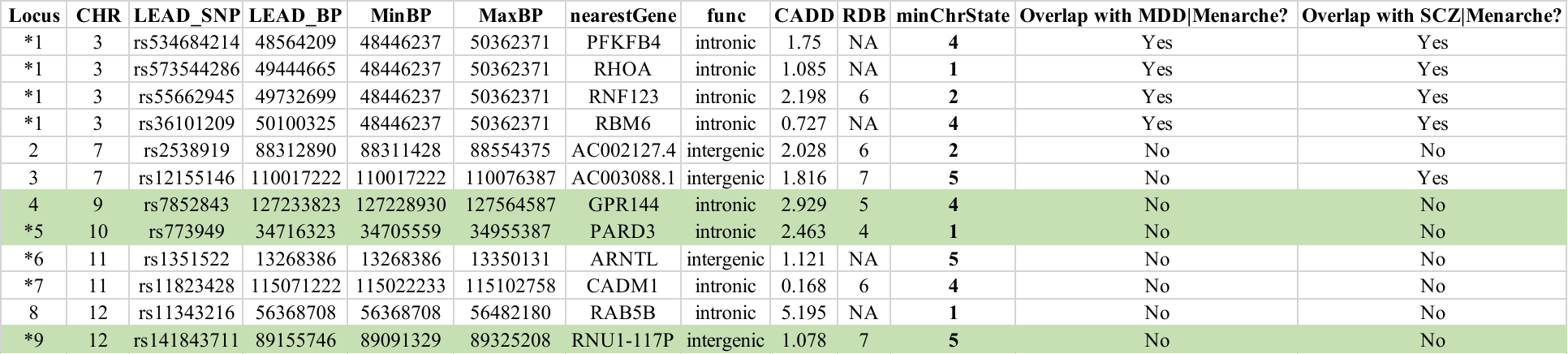

##### Supplementary Table 2: Risk loci shared between anorexia nervosa and age at menarche

Loci not previously identified as associated with Anorexia Nervosa

*loci overlapping with those identified in the conjFDR analysis of AN and body mass index

##### Supplementary Table 3: Risk loci shared between major depressive disorder and body mass index

| **Locus** | **CHR** | **LEAD_SNP** | **LEAD_BP** | **MinBP** | **MaxBP** |
| --- | --- | --- | --- | --- | --- |
| 1 | 1 | rs11121210 | 8708529 | 8457648 | 8895970 |
| 2 | 1 | rs2869511 | 23140700 | 23131185 | 23142871 |
| 3 | 1 | rs6700838 | 47700027 | 47659445 | 47708112 |
| 4 | 1 | rs6679408 | 50591565 | 49463925 | 50591851 |
| 5 | 1 | rs7535868 | 67235541 | 67225491 | 67255229 |
| 6 | 1 | rs7531118 | 72837239 | 72512988 | 73264598 |
| 7 | 1 | rs2391199 | 93160902 | 92661882 | 93440937 |
| 8 | 1 | rs6677997 | 96954561 | 96883062 | 97037083 |
| 9 | 1 | rs17496332 | 107546375 | 107537916 | 107627697 |
| 10 | 1 | rs12741940 | 175908200 | 175902596 | 176416712 |
| 11 | 1 | rs10489219 | 177326069 | 177309490 | 177428790 |
| 12 | 1 | rs2488401 | 197702401 | 197342380 | 197813558 |
| 13 | 2 | rs519111 | 25333735 | 25163783 | 25340610 |
| 14 | 2 | rs3749147 | 27851918 | 27755825 | 27967260 |
| 15 | 2 | rs1559468 | 41556924 | 41538744 | 41791788 |
| 16 | 2 | rs17041599 | 51567535 | 51503072 | 51600949 |
| 17 | 2 | rs11682175 | 57987593 | 57931347 | 58484172 |
| 18 | 2 | rs10188273 | 104209003 | 104069784 | 104479862 |
| 19 | 2 | rs16836383 | 137508441 | 137465622 | 137522867 |
| 20 | 2 | rs11695013 | 157057487 | 157014004 | 157150188 |
| 21 | 2 | rs1469015 | 160241936 | 160219552 | 160241998 |
| 22 | 2 | rs4664299 | 160570033 | 160570033 | 160667744 |
| 23 | 2 | rs6732107 | 161031369 | 160949501 | 161073157 |
| 24 | 2 | rs6738445 | 172599615 | 172521827 | 172911094 |
| 25 | 2 | rs283470 | 233656918 | 233559312 | 233763980 |
| 26 | 3 | rs10514710 | 44883971 | 44762830 | 44883971 |
| 27 | 3 | rs2681781 | 49898273 | 49734229 | 50488067 |
| 28 | 3 | rs9853056 | 52555957 | 52277445 | 53175017 |
| 29 | 3 | rs1470035 | 61184321 | 61074520 | 61277928 |
| 30 | 3 | rs4273371 | 108119071 | 108114482 | 108120008 |
| 31 | 3 | rs2693540 | 158001555 | 157829953 | 158534111 |
| 32 | 3 | rs792361 | 173044559 | 173044559 | 173127808 |
| 33 | 4 | rs4975311 | 143500223 | 143486962 | 143924210 |
| 34 | 5 | rs16903275 | 87950543 | 87936379 | 88010829 |
| 35 | 5 | rs288159 | 107364363 | 107316547 | 107684324 |
| 36 | 5 | rs10050852 | 120209016 | 120064020 | 120220251 |
| 37 | 5 | rs12515871 | 124312156 | 124295770 | 124318520 |
| 38 | 5 | rs1895336 | 144465810 | 144462121 | 144571556 |
| 39 | 5 | rs890798 | 153495869 | 153495768 | 153564109 |
| 40 | 5 | rs13178865 | 164637211 | 164465319 | 164678946 |
| 41 | 6 | rs1304602 | 40013471 | 39953541 | 40118352 |
| 42 | 6 | rs2579998 | 51477322 | 50597378 | 51481315 |
| 43 | 6 | rs1482445 | 64679917 | 64629194 | 64679917 |
| 44 | 6 | rs10457800 | 100936628 | 100890654 | 101044487 |
| 45 | 6 | rs11153066 | 107846644 | 107682044 | 107916530 |
| 46 | 6 | rs9496439 | 142993854 | 142885951 | 143004966 |
| 47 | 7 | rs12699110 | 71344395 | 71250711 | 71386881 |
| 48 | 7 | rs6964833 | 74101909 | 74027839 | 74244573 |
| 49 | 7 | rs1167827 | 75163169 | 75044830 | 75183403 |
| 50 | 7 | rs7778502 | 109101305 | 109097792 | 109144544 |
| 51 | 8 | rs13254942 | 10257678 | 10220412 | 10277982 |
| 52 | 8 | rs6997359 | 14220732 | 14176447 | 14291586 |
| 53 | 8 | rs10156227 | 76573860 | 76569119 | 76590381 |
| 54 | 9 | rs10959687 | 11280652 | 11136452 | 11887262 |
| 55 | 9 | rs13295115 | 37157964 | 37044123 | 37379492 |
| 56 | 9 | rs12553508 | 96446445 | 96386760 | 96484342 |
| 57 | 9 | rs7044210 | 126537668 | 126452936 | 126714308 |
| 58 | 9 | rs7045289 | 129913558 | 129625226 | 129948771 |
| 59 | 10 | rs1462814 | 67950100 | 67809879 | 68016799 |
| 60 | 10 | rs6480792 | 77639843 | 77537562 | 77660765 |
| 61 | 10 | rs10786634 | 103164342 | 103109066 | 103334981 |
| 62 | 10 | rs11191499 | 104764271 | 104299191 | 105059896 |
| 63 | 11 | rs7126796 | 13340867 | 13268067 | 13348783 |
| 64 | 11 | rs6416056 | 27646745 | 27457128 | 27742447 |
| 65 | 11 | rs605765 | 30217503 | 30217503 | 30395895 |
| 66 | 11 | rs3026401 | 31807524 | 31807524 | 31813529 |
| 67 | 11 | rs10768437 | 38976749 | 38807431 | 38998930 |
| 68 | 11 | rs11039024 | 46923168 | 46447246 | 48573830 |
| 69 | 11 | rs10896012 | 65278461 | 65217626 | 65343399 |
| 70 | 11 | rs7948789 | 112839532 | 112826867 | 113316102 |
| 71 | 11 | rs12273396 | 133856043 | 133707509 | 133856043 |
| 72 | 12 | rs11174358 | 62561299 | 62536274 | 62582231 |
| 73 | 13 | rs9538152 | 59227994 | 59214835 | 59300081 |
| 74 | 13 | rs1326099 | 59888054 | 59832775 | 59899311 |
| 75 | 13 | rs9599161 | 67434016 | 67414341 | 67492419 |
| 76 | 13 | rs2892657 | 94049074 | 94035548 | 94119451 |
| 77 | 13 | rs2389631 | 96932868 | 96895532 | 97029792 |
| 78 | 14 | rs17592050 | 34019770 | 34004247 | 34032771 |
| 79 | 14 | rs3007105 | 47367616 | 47156686 | 47416511 |
| 80 | 14 | rs7148764 | 75024433 | 75019157 | 75056894 |
| 81 | 14 | rs10150964 | 91507280 | 91435705 | 91549351 |
| 82 | 14 | rs11160134 | 94126515 | 94054707 | 94186395 |
| 83 | 14 | rs8010932 | 103238458 | 103229696 | 103387971 |
| 84 | 14 | rs12434218 | 104074921 | 103833065 | 104186052 |
| 85 | 15 | rs7173588 | 38910544 | 38865321 | 38925195 |
| 86 | 15 | rs9944219 | 46500612 | 46305384 | 46586095 |
| 87 | 15 | rs17648140 | 51694492 | 51683865 | 51745277 |
| 88 | 15 | rs11635675 | 63793238 | 63781143 | 63795628 |
| 89 | 15 | rs12905798 | 69551621 | 69415482 | 69597471 |
| 90 | 15 | rs4587942 | 78119345 | 78069431 | 78159256 |
| 91 | 16 | rs9928448 | 30072530 | 29924422 | 30118345 |
| 92 | 16 | rs8057044 | 53812614 | 53797908 | 53845169 |
| 93 | 16 | rs891124 | 71440756 | 71407530 | 72233195 |
| 94 | 17 | rs9299 | 46669430 | 46558036 | 46682630 |
| 95 | 17 | rs12602912 | 65870073 | 65822573 | 66096529 |
| 96 | 18 | rs11081818 | 31251088 | 31186985 | 31298923 |
| 97 | 18 | rs10502716 | 36560067 | 36513326 | 36961068 |
| 98 | 18 | rs8092503 | 52479487 | 51553172 | 52569120 |
| 99 | 18 | rs1942204 | 68714195 | 68682216 | 68714195 |
| 100 | 19 | rs12985909 | 18439383 | 18412122 | 18444809 |
| 101 | 20 | rs13043475 | 51209603 | 50912495 | 51237903 |
| 102 | 21 | rs7281757 | 46636161 | 46488169 | 46647078 |
| 103 | 22 | rs6001848 | 40636799 | 40558064 | 40722745 |
| 104 | 22 | rs926914 | 41418154 | 41215672 | 41713111 |

##### Supplementary Table 4: Risk loci shared between major depressive disorder and age at menarche

| **Locus** | **CHR** | **LEAD_SNP** | **LEAD_BP** | **MinBP** | **MaxBP** |
| --- | --- | --- | --- | --- | --- |
| 1 | 1 | rs12028339 | 7784902 | 7784658 | 7794111 |
| 2 | 1 | rs301816 | 8505058 | 8411211 | 8898807 |
| 3 | 1 | rs6689755 | 67113050 | 67022851 | 67125691 |
| 4 | 1 | rs3101336 | 72751185 | 72512988 | 72959039 |
| 5 | 1 | rs6662366 | 91230809 | 91189731 | 91234126 |
| 6 | 1 | rs2151623 | 96871770 | 96871392 | 96878813 |
| 7 | 1 | rs2759328 | 173876705 | 173539617 | 174589019 |
| 8 | 1 | rs2290754 | 243806268 | 243674682 | 244011843 |
| 9 | 2 | rs17524070 | 51565478 | 51554749 | 51666021 |
| 10 | 2 | rs13023050 | 142496643 | 142496643 | 142497249 |
| 11 | 2 | rs1534752 | 156711933 | 156614382 | 156764503 |
| 12 | 3 | rs1881915 | 44579918 | 44458730 | 44912443 |
| 13 | 3 | rs9834003 | 49216472 | 48723302 | 50250837 |
| 14 | 3 | rs17723244 | 117710096 | 117352205 | 117822657 |
| 15 | 3 | rs10935037 | 132650822 | 132622606 | 132680873 |
| 16 | 4 | rs2051559 | 3298800 | 3281369 | 3383552 |
| 17 | 5 | rs7729225 | 77142829 | 77135895 | 77157607 |
| 18 | 5 | rs768705 | 87568710 | 87513722 | 87778792 |
| 19 | 5 | rs2044318 | 133902288 | 133875033 | 133905829 |
| 20 | 5 | rs890798 | 153495869 | 153495768 | 153564109 |
| 21 | 6 | rs240766 | 100954036 | 100890654 | 101614312 |
| 22 | 6 | rs314262 | 105394621 | 105347985 | 105475254 |
| 23 | 7 | rs10257990 | 2232671 | 2171704 | 2247496 |
| 24 | 7 | rs6967152 | 74099317 | 74027839 | 74244573 |
| 25 | 7 | rs10249457 | 117652571 | 117542907 | 117672231 |
| 26 | 8 | rs7841046 | 41204825 | 41202004 | 41218282 |
| 27 | 8 | rs10156227 | 76573860 | 76569119 | 76590381 |
| 28 | 8 | rs16939338 | 77630231 | 77588548 | 77695732 |
| 29 | 8 | rs7832567 | 94628976 | 94528712 | 94651024 |
| 30 | 9 | rs10809393 | 11227070 | 11141030 | 11629615 |
| 31 | 9 | rs7044599 | 86757137 | 86744567 | 86762039 |
| 32 | 9 | rs11790449 | 96288986 | 96181075 | 96381916 |
| 33 | 9 | rs10481683 | 119803933 | 119803175 | 119803933 |
| 34 | 10 | rs1149716 | 77050387 | 77002679 | 77119918 |
| 35 | 10 | rs7893954 | 104318966 | 104222963 | 104583932 |
| 36 | 10 | rs17095748 | 118916227 | 118900107 | 118917154 |
| 37 | 11 | rs4074134 | 27647285 | 27541623 | 28543812 |
| 38 | 11 | rs1783980 | 57448325 | 57409538 | 57682011 |
| 39 | 12 | rs12425203 | 2474661 | 2285731 | 2523772 |
| 40 | 12 | rs10783486 | 52362786 | 52343231 | 52396241 |
| 41 | 12 | rs668622 | 121198299 | 121187557 | 121378596 |
| 42 | 12 | rs7132277 | 123593382 | 123450765 | 123913433 |
| 43 | 13 | rs1327958 | 59843978 | 59832775 | 59889839 |
| 44 | 14 | rs3008494 | 85013821 | 84908095 | 85035235 |
| 45 | 14 | rs17101957 | 104410305 | 104410305 | 104511206 |
| 46 | 15 | rs6493198 | 46493154 | 46305384 | 46618157 |
| 47 | 15 | rs8040348 | 47999319 | 47982997 | 48083383 |
| 48 | 16 | rs9928448 | 30072530 | 29924422 | 30118345 |
| 49 | 16 | rs8057044 | 53812614 | 53797908 | 53845169 |
| 50 | 17 | rs7223364 | 7780709 | 7750232 | 7785257 |
| 51 | 17 | rs1285293 | 77924776 | 77917191 | 77926491 |
| 52 | 19 | rs17674370 | 3295428 | 3274770 | 3300401 |
| 53 | 20 | rs293559 | 31089109 | 30702144 | 31091206 |
| 54 | 20 | rs2273653 | 47770756 | 47511792 | 47924894 |

##### Supplementary Table 5: Risk loci shared between schizophrenia and body mass index

| **Locus** | **CHR** | **LEAD_SNP** | **LEAD_BP** | **MinBP** | **MaxBP** |
| --- | --- | --- | --- | --- | --- |
| 1 | 1 | rs6577584 | 6715390 | 6644723 | 6788985 |
| 2 | 1 | rs2016084 | 8613418 | 8431607 | 8892577 |
| 3 | 1 | rs6694677 | 41058237 | 41055384 | 41058237 |
| 4 | 1 | rs6679408 | 50591565 | 49239968 | 50591851 |
| 5 | 1 | rs2590942 | 72885281 | 72628347 | 72959039 |
| 6 | 1 | rs1409058 | 97106177 | 97075059 | 97112259 |
| 7 | 1 | rs1218582 | 154834183 | 154834092 | 154919565 |
| 8 | 1 | rs10489219 | 177326069 | 177309490 | 177428790 |
| 9 | 1 | rs1410402 | 193648980 | 193576625 | 193699448 |
| 10 | 1 | rs2242000 | 205031769 | 205023028 | 205173866 |
| 11 | 1 | rs12744297 | 243734296 | 243690491 | 244025317 |
| 12 | 2 | rs7603056 | 27064608 | 26961166 | 27119655 |
| 13 | 2 | rs6545145 | 50245211 | 50243779 | 50268711 |
| 14 | 2 | rs6714450 | 54252825 | 53864624 | 54347902 |
| 15 | 2 | rs1106090 | 58068741 | 57917222 | 58505679 |
| 16 | 2 | rs1861411 | 58904177 | 58866584 | 59039998 |
| 17 | 2 | rs6432565 | 160670424 | 160397825 | 160675650 |
| 18 | 2 | rs6738445 | 172599615 | 172521827 | 172911094 |
| 19 | 2 | rs1480481 | 185547462 | 185532918 | 185786030 |
| 20 | 2 | rs13400686 | 200289152 | 199904222 | 200313235 |
| 21 | 2 | rs16825005 | 212304565 | 212264642 | 212305124 |
| 22 | 2 | rs6435711 | 213410065 | 213401874 | 213547305 |
| 23 | 2 | rs6714784 | 215263165 | 215247715 | 215406636 |
| 24 | 2 | rs10175063 | 228980113 | 228971784 | 229017931 |
| 25 | 2 | rs283470 | 233656918 | 233559312 | 233763980 |
| 26 | 3 | rs2251361 | 9428270 | 9404877 | 9436472 |
| 27 | 3 | rs11128818 | 17143621 | 16973762 | 17151126 |
| 28 | 3 | rs6765484 | 50041313 | 49897830 | 50209053 |
| 29 | 3 | rs2710323 | 52815905 | 52536514 | 53175017 |
| 30 | 3 | rs1155530 | 59727802 | 59705135 | 59736299 |
| 31 | 3 | rs6807748 | 61149967 | 61074520 | 61277928 |
| 32 | 3 | rs17350402 | 69401498 | 69382821 | 69422357 |
| 33 | 3 | rs12485775 | 70561018 | 70472539 | 70593081 |
| 34 | 3 | rs10511111 | 80599757 | 80450582 | 81050602 |
| 35 | 3 | rs17023388 | 85877180 | 85793588 | 85880011 |
| 36 | 3 | rs11917750 | 107894492 | 107828001 | 107942785 |
| 37 | 3 | rs2903971 | 116935744 | 116925212 | 116951842 |
| 38 | 3 | rs687339 | 135932359 | 135619585 | 136671504 |
| 39 | 3 | rs9810292 | 180714307 | 180524764 | 181031667 |
| 40 | 3 | rs9831938 | 185785996 | 185733707 | 185824903 |
| 41 | 4 | rs1849338 | 45142805 | 45037953 | 45186832 |
| 42 | 4 | rs2904221 | 77182033 | 77145983 | 77188613 |
| 43 | 4 | rs13107325 | 103188709 | 102702364 | 104129141 |
| 44 | 4 | rs1992418 | 143737159 | 143629150 | 143924210 |
| 45 | 4 | rs2897478 | 153073562 | 153016290 | 153095125 |
| 46 | 5 | rs13186194 | 60795485 | 60563907 | 60844213 |
| 47 | 5 | rs16872526 | 74675717 | 74641707 | 75016943 |
| 48 | 5 | rs661311 | 88034867 | 87986239 | 88191635 |
| 49 | 5 | rs11743085 | 88825791 | 88489418 | 88854539 |
| 50 | 5 | rs10463981 | 101787062 | 101584734 | 102028537 |
| 51 | 5 | rs13178589 | 106861387 | 106855830 | 106861387 |
| 52 | 5 | rs10040792 | 137759010 | 137608056 | 137948140 |
| 53 | 5 | rs13174863 | 139080745 | 139037130 | 139086651 |
| 54 | 5 | rs12659802 | 152274478 | 151924995 | 152339648 |
| 55 | 5 | rs815609 | 153516901 | 153360230 | 153564678 |
| 56 | 5 | rs10052410 | 155714344 | 155681868 | 155727958 |
| 57 | 6 | rs10946865 | 26878864 | 26678512 | 27017827 |
| 58 | 6 | rs3828783 | 33767727 | 33513968 | 33803752 |
| 59 | 6 | rs1723535 | 64136886 | 64126180 | 64251656 |
| 60 | 6 | rs2153960 | 108988184 | 108861264 | 109019323 |
| 61 | 6 | rs1775613 | 119346010 | 119310119 | 119353212 |
| 62 | 6 | rs12194023 | 163816903 | 163792420 | 164005951 |
| 63 | 7 | rs7811417 | 21534152 | 21470536 | 21552995 |
| 64 | 7 | rs10267684 | 71626255 | 71437062 | 71872935 |
| 65 | 7 | rs7795893 | 104568299 | 104524092 | 105040962 |
| 66 | 7 | rs13246624 | 113687707 | 113349793 | 113708893 |
| 67 | 7 | rs1593312 | 131584453 | 131528516 | 131627573 |
| 68 | 8 | rs13254942 | 10257678 | 9793601 | 10283602 |
| 69 | 8 | rs919494 | 27313085 | 27152497 | 27317445 |
| 70 | 8 | rs6991355 | 34350971 | 34111195 | 34524021 |
| 71 | 8 | rs2178946 | 116537058 | 116464988 | 117132729 |
| 72 | 8 | rs6583611 | 143330576 | 143297312 | 143404118 |
| 73 | 9 | rs10511914 | 34107232 | 34081235 | 34149119 |
| 74 | 9 | rs2790063 | 37287405 | 37044388 | 37379492 |
| 75 | 9 | rs1757961 | 72068932 | 72042193 | 72166573 |
| 76 | 9 | rs11138311 | 82229981 | 82166319 | 82305000 |
| 77 | 9 | rs2997922 | 131580744 | 131561110 | 131652063 |
| 78 | 10 | rs4881164 | 3415022 | 3355368 | 3480163 |
| 79 | 10 | rs1010826 | 33473620 | 33470023 | 33518975 |
| 80 | 10 | rs10827283 | 34064066 | 33857195 | 34064066 |
| 81 | 10 | rs7068132 | 77362061 | 77325146 | 77362061 |
| 82 | 10 | rs11191560 | 104869038 | 104214048 | 105059896 |
| 83 | 11 | rs2051773 | 17365209 | 17067849 | 17421886 |
| 84 | 11 | rs879048 | 27638934 | 27541623 | 27742447 |
| 85 | 11 | rs607987 | 30223574 | 30166096 | 30395895 |
| 86 | 11 | rs12418412 | 34072011 | 34028081 | 34074411 |
| 87 | 11 | rs10768437 | 38976749 | 38807431 | 38998930 |
| 88 | 11 | rs7128102 | 46928455 | 46330604 | 47989003 |
| 89 | 11 | rs562664 | 63823619 | 63805335 | 63838452 |
| 90 | 11 | rs10502165 | 112934495 | 112826311 | 113270160 |
| 91 | 11 | rs7925214 | 130794253 | 130794253 | 130892029 |
| 92 | 11 | rs3016500 | 132581927 | 132541038 | 132581927 |
| 93 | 11 | rs329651 | 133767622 | 133707509 | 133814468 |
| 94 | 11 | rs2155296 | 134553798 | 134528720 | 134601012 |
| 95 | 12 | rs1545319 | 23537708 | 23350235 | 23637351 |
| 96 | 12 | rs7976793 | 54073069 | 54015572 | 54092021 |
| 97 | 12 | rs17464848 | 89935496 | 89799488 | 89935496 |
| 98 | 12 | rs2731283 | 90593548 | 90589436 | 90704972 |
| 99 | 12 | rs2372716 | 99573426 | 99436519 | 99691448 |
| 100 | 12 | rs1502337 | 111062852 | 110513682 | 111212762 |
| 101 | 12 | rs1063843 | 121681687 | 121639657 | 121690555 |
| 102 | 12 | rs7137049 | 122493384 | 122483836 | 123144293 |
| 103 | 12 | rs7307277 | 124475156 | 124399155 | 124500725 |
| 104 | 13 | rs3105038 | 55968795 | 55679499 | 56303709 |
| 105 | 13 | rs953618 | 57112724 | 56820837 | 57112724 |
| 106 | 13 | rs1537475 | 59229871 | 59175727 | 59300081 |
| 107 | 13 | rs338763 | 89132137 | 89060548 | 89136916 |
| 108 | 13 | rs9554338 | 96966314 | 96901225 | 97029792 |
| 109 | 13 | rs1536053 | 111982291 | 111978920 | 111982291 |
| 110 | 14 | rs1950709 | 29735938 | 29716334 | 29778184 |
| 111 | 14 | rs225884 | 30473054 | 30471987 | 30514667 |
| 112 | 14 | rs17522122 | 33302882 | 33292743 | 33309495 |
| 113 | 14 | rs3850422 | 99671788 | 99657227 | 99706474 |
| 114 | 14 | rs4906270 | 103364737 | 103251003 | 103387971 |
| 115 | 14 | rs3759579 | 103851272 | 103851272 | 104010198 |
| 116 | 15 | rs1077421 | 44236115 | 43588439 | 44443301 |
| 117 | 15 | rs1559677 | 47738063 | 47738063 | 47751837 |
| 118 | 15 | rs4774315 | 59202598 | 59136688 | 59272096 |
| 119 | 15 | rs8042567 | 67975599 | 67716572 | 68143468 |
| 120 | 15 | rs2301249 | 75092384 | 75031521 | 75222415 |
| 121 | 15 | rs12914385 | 78898723 | 78801394 | 79074518 |
| 122 | 15 | rs13380104 | 79378821 | 79372413 | 79472522 |
| 123 | 16 | rs11641136 | 19521712 | 19395409 | 19630843 |
| 124 | 16 | rs4787491 | 30015337 | 29923510 | 30343236 |
| 125 | 16 | rs1123072 | 67898797 | 67351696 | 68299697 |
| 126 | 16 | rs5475 | 72094348 | 71649997 | 72895614 |
| 127 | 16 | rs1019959 | 82431694 | 82431694 | 82438337 |
| 128 | 16 | rs455527 | 89644001 | 89593976 | 89661807 |
| 129 | 17 | rs2281727 | 2117945 | 2001825 | 2220496 |
| 130 | 17 | rs2309399 | 5391690 | 5170870 | 5415684 |
| 131 | 17 | rs11656775 | 17654319 | 17645951 | 18035019 |
| 132 | 17 | rs9899357 | 19800549 | 19799698 | 19900168 |
| 133 | 17 | rs11263770 | 34896877 | 34825861 | 34961772 |
| 134 | 17 | rs4459609 | 61548948 | 61545779 | 61579612 |
| 135 | 17 | rs11653272 | 78639760 | 78518327 | 78706517 |
| 136 | 18 | rs10401120 | 53192498 | 53155002 | 53465817 |
| 137 | 19 | rs2965185 | 19525792 | 19350103 | 19725456 |
| 138 | 19 | rs7249968 | 30711242 | 30698839 | 30718774 |
| 139 | 19 | rs11669242 | 31087575 | 31087575 | 31087575 |
| 140 | 19 | rs281377 | 49206603 | 49201217 | 49250657 |
| 141 | 20 | rs2236270 | 33523155 | 32514061 | 33536585 |
| 142 | 20 | rs734784 | 43723627 | 43723627 | 43773168 |
| 143 | 20 | rs310618 | 62127121 | 62127121 | 62129566 |
| 144 | 21 | rs2298450 | 37623447 | 37610551 | 37643609 |
| 145 | 21 | rs2256553 | 40295090 | 40290503 | 40296411 |
| 146 | 22 | rs717121 | 22081573 | 22069757 | 22093119 |
| 147 | 22 | rs926914 | 41418154 | 41215672 | 41713111 |

##### Supplementary Table 6: Risk loci shared between schizophrenia and age at menarche

| **Locus** | **CHR** | **LEAD_SNP** | **LEAD_BP** | **MinBP** | **MaxBP** |
| --- | --- | --- | --- | --- | --- |
| 1 | 1 | rs301798 | 8488565 | 8411211 | 8888842 |
| 2 | 1 | rs489319 | 44131794 | 43858630 | 44383914 |
| 3 | 1 | rs782240 | 72906385 | 72628347 | 72959039 |
| 4 | 1 | rs3856224 | 91223871 | 91219359 | 91234126 |
| 5 | 1 | rs4423049 | 98409303 | 98298371 | 98651987 |
| 6 | 1 | rs10798302 | 173987798 | 173867252 | 174643725 |
| 7 | 1 | rs10754807 | 243804158 | 243649330 | 244025317 |
| 8 | 2 | rs4618000 | 30145427 | 30132986 | 30145427 |
| 9 | 2 | rs988911 | 61607510 | 61367664 | 61782899 |
| 10 | 2 | rs3963519 | 200007302 | 199688708 | 200347991 |
| 11 | 2 | rs6709782 | 236770075 | 236769459 | 236827263 |
| 12 | 3 | rs2624843 | 49997963 | 48446237 | 50228316 |
| 13 | 3 | rs4677034 | 71368262 | 71362196 | 71407890 |
| 14 | 3 | rs6769923 | 107205050 | 107169235 | 107228565 |
| 15 | 3 | rs861001 | 117720355 | 117389497 | 117820386 |
| 16 | 3 | rs2999060 | 127900494 | 127710474 | 128122396 |
| 17 | 3 | rs16860342 | 185652947 | 185623175 | 185824903 |
| 18 | 4 | rs4697446 | 24269622 | 24264014 | 24270210 |
| 19 | 4 | rs12648749 | 28713799 | 28575886 | 28831239 |
| 20 | 4 | rs1849338 | 45142805 | 45068929 | 45186832 |
| 21 | 4 | rs6816787 | 102795845 | 102766892 | 102849219 |
| 22 | 5 | rs4451795 | 87718696 | 87513775 | 87807553 |
| 23 | 5 | rs10055995 | 137698299 | 137585817 | 137837245 |
| 24 | 5 | rs319234 | 146241232 | 146229816 | 146337743 |
| 25 | 5 | rs12054772 | 153509357 | 153360230 | 153577521 |
| 26 | 6 | rs9470808 | 12035486 | 11957303 | 12038979 |
| 27 | 6 | rs3999543 | 26259807 | 26180634 | 26336300 |
| 28 | 6 | rs9461366 | 27310533 | 27304586 | 27310533 |
| 29 | 6 | rs213236 | 28324397 | 27895213 | 28668072 |
| 30 | 6 | rs314262 | 105394621 | 105347985 | 105475254 |
| 31 | 6 | rs11154433 | 128314604 | 128301981 | 128333682 |
| 32 | 7 | rs3778969 | 2139990 | 1909865 | 2247496 |
| 33 | 7 | rs1476665 | 41706132 | 41706132 | 41743364 |
| 34 | 7 | rs11764402 | 110037249 | 110017222 | 110076387 |
| 35 | 8 | rs16881274 | 33713382 | 33551744 | 33968227 |
| 36 | 8 | rs12547436 | 40747667 | 40712159 | 40771375 |
| 37 | 8 | rs747498 | 131030580 | 131027338 | 131165086 |
| 38 | 8 | rs13252406 | 144912303 | 144870701 | 144933453 |
| 39 | 9 | rs913589 | 7174773 | 7164873 | 7200125 |
| 40 | 9 | rs10217711 | 10257826 | 10222902 | 10277582 |
| 41 | 9 | rs7874413 | 22818289 | 22765073 | 22889446 |
| 42 | 9 | rs1180117 | 72897440 | 72749590 | 72984008 |
| 43 | 9 | rs10780534 | 84376404 | 84294425 | 84384450 |
| 44 | 9 | rs10821136 | 96238731 | 96161300 | 96381916 |
| 45 | 9 | rs2289480 | 111705501 | 111661422 | 111879398 |
| 46 | 10 | rs7893954 | 104318966 | 104318966 | 104318966 |
| 47 | 10 | rs2066323 | 104871361 | 104595420 | 104962011 |
| 48 | 10 | rs7923863 | 123717399 | 123579187 | 123729960 |
| 49 | 11 | rs879048 | 27638934 | 27541623 | 27742447 |
| 50 | 11 | rs7947147 | 28425239 | 27996573 | 28577867 |
| 51 | 11 | rs7476 | 46342834 | 45949201 | 46751495 |
| 52 | 11 | rs1783980 | 57448325 | 57409538 | 57682011 |
| 53 | 11 | rs570098 | 63787389 | 63585804 | 63797679 |
| 54 | 12 | rs4441076 | 2502058 | 2285731 | 2523772 |
| 55 | 12 | rs6606711 | 109849297 | 109788019 | 110027795 |
| 56 | 12 | rs7132277 | 123593382 | 123450765 | 123913433 |
| 57 | 13 | rs949466 | 56510762 | 56438043 | 56825218 |
| 58 | 14 | rs17129021 | 93819544 | 93819216 | 94032195 |
| 59 | 14 | rs1152790 | 99710843 | 99707933 | 99718105 |
| 60 | 15 | rs8042567 | 67975599 | 67716572 | 68143468 |
| 61 | 15 | rs17484524 | 78772676 | 78719501 | 78912710 |
| 62 | 15 | rs2099259 | 83323688 | 82827938 | 83406857 |
| 63 | 16 | rs3809624 | 30102802 | 29923510 | 30430571 |
| 64 | 17 | rs1602914 | 5190108 | 5156950 | 5415684 |
| 65 | 17 | rs7502667 | 71894357 | 71875014 | 71929602 |
| 66 | 17 | rs7501740 | 78610345 | 78456708 | 78704618 |
| 67 | 19 | rs6511046 | 19727042 | 19668738 | 19753292 |
| 68 | 19 | rs10518269 | 31028666 | 30982165 | 31051857 |
| 69 | 19 | rs281379 | 49214274 | 49168942 | 49252574 |
| 70 | 20 | rs220498 | 37297860 | 37266561 | 37298929 |
| 71 | 20 | rs2267850 | 43524963 | 43503276 | 43544528 |
| 72 | 20 | rs6012555 | 47527742 | 47511792 | 47914180 |
| 73 | 21 | rs7276774 | 40628725 | 40513750 | 40709960 |

##### Supplementary Table 7: AN-associated risk loci overlapping with different evolutionary methods

| ***A) Loci overlapping with regions with the 5% lowest NSS^(4)^*** | | | | | | | |
| --- | --- | --- | --- | --- | --- | --- | --- |
| **Chr** | **Lead SNPs** | **Lead BP** | **MinBP** | **MaxBP** | **nearestGene** | **AN analysis** | **Additional information** |
| 1 | rs4000303 | 96913765 | 96883812 | 97296935 | *EEF1A1P11* | AN+BMI | this region is located at the *PTBP2* gene locus |
| 1 | rs1236381 | 97005643 | 96883812 | 97296935 | *RN7SL831P* | AN+BMI |  |
| 1 | rs10747478 | 96901455 | 96883812 | 96972973 |  | AN main GWAS |  |
| 3 | rs534684214 | 48564209 | 48446237 | 50362371 | *PFKFB4* | AN+MENA | this cover two regions with the 5% smallest NSS, one with a mean NSS of -4.67 (covering *TMEM89; SLC26A6, CELSR3, NCKIPSD, IP6K2, PRKAR2A*) and one with a mean NSS of -6.02 (including the *QRICH11, QARS1, USP19, LAMB2, IHO1, CCDC71, KLHDC8B, USP4, RHOA,GPX1* gene locus) |
| 3 | rs573544286 | 49444665 | 48446237 | 50362371 | *RHOA* | AN+MENA |  |
| 3 | rs55662945 | 49732699 | 48446237 | 50362371 | *RNF123* | AN+MENA |  |
| 3 | rs36101209 | 50100325 | 48446237 | 50362371 | *RBM6* | AN+MENA |  |
| 3 | rs9821797 | 48718253 | 48446237 | 50552866 |  | AN main GWAS |  |
| 3 | rs73080977 | 50189531 | 48446237 | 50552866 |  | AN main GWAS |  |
| 3 | rs9873183 | 49734229 | 49734229 | 50209053 | *RNF123* | AN+BMI |  |
| 3 | rs13100344 | 94605107 | 94520179 | 94738138 |  | AN main GWAS | intergenic region close to *NSUN3, ARL13B, STX19, DHFR2, DHFRL1, PROS1* |
| 12 | rs141843711 | 89155746 | 89091329 | 89411738 | *RNU1-117P* | AN+BMI | intergenic region close to *KITLG* |
| 12 | rs141843711 | 89155746 | 89091329 | 89325208 | *RNU1-117P* | AN+MENA |  |
| ***B) Loci overlapping (+/- 10kb) with regions identified by the Composite of Multiple Signal method^(5)^*** | | | | | | | |
| **Chr** | **Lead SNPs** | **Lead BP** | **MinBP** | **MaxBP** | **nearestGene** | **AN analysis** | **Additional information** |
| 3 | rs534684214 | 48564209 | 48446237 | 50362371 | PFKFB4 | AN+MENA | region 66 and region 112 from CMS |
| 3 | rs573544286 | 49444665 | 48446237 | 50362371 | RHOA | AN+MENA |  |
| 3 | rs55662945 | 49732699 | 48446237 | 50362371 | RNF123 | AN+MENA |  |
| 3 | rs36101209 | 50100325 | 48446237 | 50362371 | RBM6 | AN+MENA |  |
| 3 | rs9821797 | 48718253 | 48446237 | 50552866 |  | AN main GWAS |  |
| 3 | rs73080977 | 50189531 | 48446237 | 50552866 |  | AN main GWAS |  |
| 3 | rs9873183 | 49734229 | 49734229 | 50209053 | RNF123 | AN+BMI |  |
| 12 | rs141843711 | 89155746 | 89091329 | 89411738 | RNU1-117P | AN+BMI | region at the *KILTG* gene locus |
| 12 | rs141843711 | 89155746 | 89091329 | 89325208 | RNU1-117P | AN+MENA |  |
| ***C) Loci overlapping (+/- 10kb) with regions identified as differently methylated between humans and neanderthal^(6)^*** | | | | | | | |
| **Chr** | **Lead SNPs** | **Lead BP** | **MinBP** | **MaxBP** | **nearestGene** | **AN analysis** | **Additional information** |
| 1 | rs72634839 | 1674111 | 1658343 | 1879958 | SLC35E2 | AN+BMI | Differently methylation region in the promoter of *CALML6* (93,6% in present-day human osteoblast vs 33,0% in Neanderthal) |
| 3 | rs534684214 | 48564209 | 48446237 | 50362371 | PFKFB4 | AN+MENA | Overlap with three differently methylation regions: a) one within a CpG island in the promoter of *MST1R* (6,2% in present-day human osteoblast vs 76,5% in Neanderthal); b) one in the gene body of *SEMA3F* (37,3% in present-day human osteoblast vs 100,0% in Neanderthal); and c) one intergenic region (10,8% in present-day human osteoblast and 72,4% in Neanderthal) |
| 3 | rs573544286 | 49444665 | 48446237 | 50362371 | RHOA | AN+MENA |  |
| 3 | rs55662945 | 49732699 | 48446237 | 50362371 | RNF123 | AN+MENA |  |
| 3 | rs36101209 | 50100325 | 48446237 | 50362371 | RBM6 | AN+MENA |  |
| 3 | rs9821797 | 48718253 | 48446237 | 50552866 |  | AN main GWAS |  |
| 3 | rs73080977 | 50189531 | 48446237 | 50552866 |  | AN main GWAS |  |
| 3 | rs9873183 | 49734229 | 49734229 | 50209053 | RNF123 | AN+BMI |  |

Locus highlighted by all three evolutionary methods
